# Longitudinal study of DNA methylation and epigenetic clocks prior to and following test-confirmed COVID-19 and mRNA vaccination

**DOI:** 10.1101/2021.12.01.21266670

**Authors:** Alina PS Pang, Albert T. Higgins-Chen, Florence Comite, Ioana Raica, Christopher Arboleda, Hannah Went, Tavis Mendez, Michael Schotsaert, Varun Dwaraka, Ryan Smith, Morgan E. Levine, Lishomwa C. Ndhlovu, Michael J. Corley

## Abstract

The host epigenetic landscape is rapidly changed during SARS-CoV-2 infection and evidence suggests that severe COVID-19 is associated with durable scars to the epigenome. Specifically, aberrant DNA methylation changes in immune cells and alterations to epigenetic clocks in blood relate to severe COVID-19. However, a longitudinal assessment of DNA methylation states and epigenetic clocks in blood from healthy individuals prior to and following test-confirmed non-hospitalized COVID-19 has not been performed. Moreover, the impact of mRNA COVID-19 vaccines upon the host epigenome remains understudied. Here, we first examined DNA methylation states in blood of 21 participants prior to and following test confirmed COVID-19 diagnosis at a median timeframe of 8.35 weeks. 261 CpGs were identified as differentially methylated following COVID-19 diagnosis in blood at an FDR adjusted P value <0.05. These CpGs were enriched in gene body and northern and southern shelf regions of genes involved in metabolic pathways. Integrative analysis revealed overlap among genes identified in transcriptional SARS-CoV-2 infection datasets. Principal component-based epigenetic clock estimates of PhenoAge and GrimAge significantly increased in people over 50 following infection by an average of 2.1 and 0.84 years. In contrast, PCPhenoAge significantly decreased in people under 50 following infection by an average of 2.06 years. This observed divergence in epigenetic clocks following COVID-19 was related to age and immune cell-type compositional changes in CD4+ T cells, B cells, granulocytes, plasmablasts, exhausted T cells, and naïve T cells. Complementary longitudinal epigenetic clock analyses of 36 participants prior to and following Pfizer and Moderna mRNA-based COVID-19 vaccination revealed vaccination significantly reduced principal component-based Horvath epigenetic clock estimates in people over 50 by an average of 3.91 years for those that received Moderna. This reduction in epigenetic clock estimates was significantly related to chronological age and immune cell-type compositional changes in B cells and plasmablasts pre- and post-vaccination. These findings suggest the potential utility of epigenetic clocks as a biomarker of COVID-19 vaccine responses. Future research will need to unravel the significance and durability of short-term changes in epigenetic age related to COVID-19 exposure and mRNA vaccination.

## 1 Introduction

Epigenetic mechanisms including DNA methylation are critically involved in both host immune responses to viral infection and subsequent diseases pathogenesis and severity (Gómez-Díaz et al., 2012; Morales-Nebreda et al., 2019). In the context of SARS-CoV-2 infection, human studies suggest that DNA methylation states in immune cells are altered during infection and associate with COVID-19 disease severity (Bernardes et al., 2020; Balnis et al., 2021; Castro de Moura et al., 2021; Corley et al., 2021). We previously reported a unique candidate immune cell DNA methylation signature associated with severe COVID-19 that was distinct from influenza, primary HIV infection, and HIV/mild COVID-19 coinfection (Corley et al., 2021). Additional studies have extended these findings and reported distinct genome-wide DNA methylation differences in peripheral blood from COVID-19 patients based on disease severity (Balnis et al., 2021; Castro de Moura et al., 2021; Zhou et al., 2021). Insights into host DNA methylation states and COVID-19 have mainly focused on severe COVID-19 and are limited by the use of cross-sectional study designs. Longitudinal epigenetic studies of COVID-19 are lacking, and it remains unclear whether rapid changes to immune cell epigenetic DNA methylation patterns occur in healthy individuals that recover from non-hospitalized COVID-19.

The severity of COVID-19 strongly depends on age, and aging biomarkers may help explain this relationship and predict who is at highest risk of severe COVID-19 (Kuo et al., 2020; Mueller et al., 2020). Distinct DNA methylation patterns have been utilized to derive epigenetic measures of biological aging termed “epigenetic clocks”(Horvath, 2013). Numerous epigenetic clocks have been generated that appear to capture distinct aspects of aging and associate with different biological hallmarks of aging, environmental exposures, traits, and disease patterns (Horvath and Raj, 2018; Liu et al., 2020; Higgins-Chen et al., 2021a; Oblak et al., 2021). Many of the differences between clocks stem from being trained to predict different aging-related variables, such as chronological age, mortality risk, or mitotic divisions. Moreover, epigenetic clocks are accurate predictors of mortality risk (Lu et al., 2019), biomarkers of pathogen exposure (Horvath and Levine, 2015; Boulias et al., 2016; Corley et al., 2021), and correlates of lung function and immune inflammation (Hillary et al., 2020, 2021). Evidence suggests that severe COVID-19 disease may impact certain epigenetic clocks (Corley et al., 2021; Mongelli et al., 2021a) and biological aging captured by PhenoAge may inform COVID-19 outcomes (Kuo et al., 2020). More recent epigenetic clock studies have reported conflicting evidence for biological age acceleration and telomere shortening in COVID-19 survivors (Mongelli et al., 2021b), with some finding no clock acceleration in COVID-19 patients (Franzen et al., 2021). Whether changes occur to epigenetic clocks in healthy individuals that recover from non-hospitalized COVID-19 remains unclear. Additionally, whether epigenetic clocks are impacted following mRNA COVID-19 vaccination remains understudied.

In this study, we first examined whether alterations to DNA methylation states, blood immune cell type composition, and epigenetic clocks occurred in peripheral blood following COVID-19 using a longitudinal study design of 21 healthy participants prior to and following test-confirmed COVID-19. Next, we also evaluated longitudinal DNA methylation states, blood immune cell type composition, and epigenetic clocks of 36 healthy participants prior to and following complete two-dose mRNA-based COVID-19 vaccination.

## 2 Results

### 2.1 Cohort of participants with longitudinal assessments of DNA methylation prior to and following test-confirmed COVID-19 infection

**Table 1** presents the baseline (pre-COVID-19) characteristics of study participants prior to COVID-19 diagnosis. Participants were healthy (n=14M, 7F) and ranged in chronological age from 18 to 73 years (Median = 46 years). Genome-wide DNA methylation was assayed from blood biospecimens of all participants at baseline and post-COVID-19 using the Illumina MethylationEPIC platform (Pidsley et al., 2016). Baseline DNA methylation for participants was obtained at a median of 19 weeks prior to the first COVID-19-positive test (Range: 4 - 50 weeks). COVID-19 exposure and SARS-CoV-2 infection of participants for the post-COVID-19 timepoint was confirmed utilizing clinical PCR testing (n=18) and serology testing (n=3). Post-COVID-19 DNA methylation was assessed at a median timeframe of 8.35 weeks after testing positive. The earliest captured participant’s post-COVID-19 DNA methylation was within 1 week following COVID-19 diagnosis and ranged out to a maximum of 6 months after diagnosis.

**Table 1.**

|  | Pre-COVID-19 | Post-COVID-19 |
| --- | --- | --- |
| Age (year)<sup>a</sup> | 46.07 (18.53, 73.03) | 46.54 (19.41, 73.66) |
| Sex (male, %) | 14 (66.67%) | - |
| Time before COVID-19 Diagnosis DNAm assayed (Weeks) | 19.39 (4.35, 49.83) | - |
| Time after COVID-19 Diagnosis DNAm assayed (Weeks) | - | 8.35 (1.00, 27.10) |
| PCR Test Confirmed (%) | - | 85.71% |
| Antibody Test Confirmed (%) | - | 14.29% |

<sup>a</sup>Data are median (Minimum, Maximum)

### 2.2 Differentially methylated loci following SARS-CoV-2 infection

To identify differentially methylated loci in blood related to COVID-19, we utilized a longitudinal study design that included genome-wide DNA methylation data generated from 21 participants prior to (pre-COVID-19) and following COVID-19 diagnosis (post-COVID-19) (**Fig.1a**). Our repeated measures analysis of DNA methylation at pre- and post-COVID-19 timepoints revealed 756 differentially methylated loci significant at FDR (Benjamini-Hochberg) adjusted *P* < 0.05 that were not significantly biased to a specific chromosomal location (**Fig.1b**, **Supplementary Dataset 1**). 57.8% of the COVID-19 related DML were increases in DNA methylation states (hyper-methylation) at the post-COVID-19 compared to pre-timepoint for participants. Next, we examined whether these COVID-19 related DML in blood were enriched in specific genomic contexts and found a significant enrichment in gene body (Odds Ratio= 1.2 ; *P* = 0.005) and northern (Odds Ratio= 1.7 ; *P* = 0.0004) and southern (Odds Ratio= 1.8 ; *P* = 0.0001) shelf regions located adjacent to CpG island shore regions compared to the expected distribution of methylation sites assayed across the human genome (**Fig.1c**), suggestive of perturbations at regulatory regions in the human genome likely linked to transcriptional differences. We observed that the 756 COVID-19 associated DML were related to 516 annotated protein coding genes (**Supplementary Dataset 1)**. Gene enrichment analysis revealed the top biological processes involved cellular glucose homeostasis (GO: 0001678; *P* = 0.001) **Supplementary Figure S1**. KEGG pathway analyses showed the top pathway involved thyroid hormone signaling (*P* = 0.00001) **Supplementary Figure S1**. These findings support the interplay between host metabolism and metabolic gene pathways in COVID-19 and reports of dysregulated glycemia in COVID-19 (Reiterer et al., 2021).

**Figure 1.**
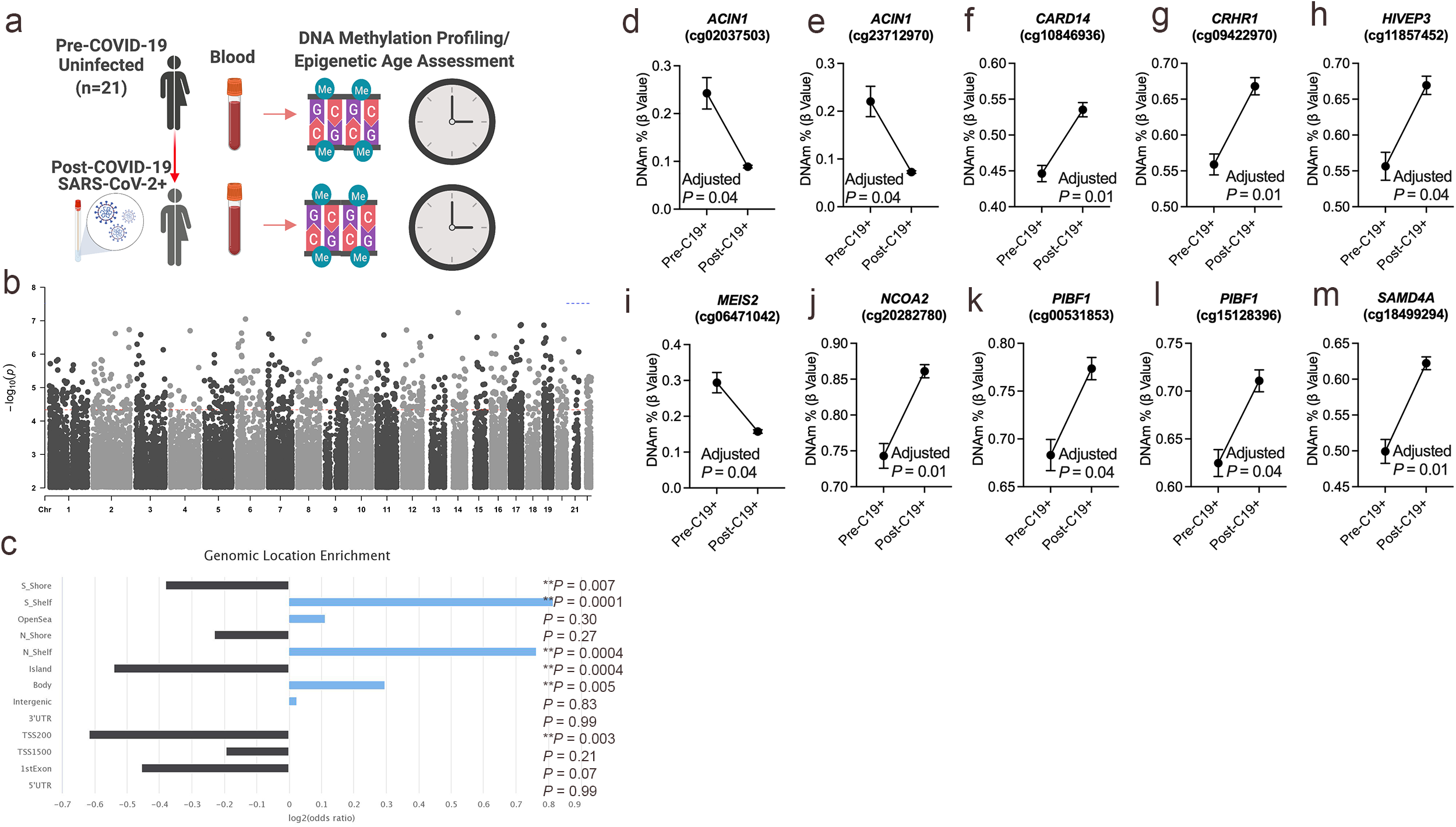
DNA methylation changes in blood associated with mild/moderate COVID-19. **a.** Study design of longitudinal assessment of DNA methylation profiles in 21 participants pre- and post-SARS-CoV-2 infection. **b.** Manhattan plot of differentially methylated loci (DML) associated with mild/moderate COVID-19. **c.** Bar graph of genomic enrichment of COVID-19 DML in 13 different categorized regions of the genome relative to gene and CpG island. Hypergeometric test utilized to calculate P value and odd ratio. **d-m.** Plots of COVID-19 DML displaying mean DNA methylation levels +/- SEM for CpGs associated with a gene ID. Adjusted P value calculated utilizing Benjamini-Hochberg correction.

Since our assessments of DNA methylation in participants occurred in whole blood, we next applied a bioinformatic tool to identify the potential cellular source in blood of the COVID-19 exposure-related differences in participants. We analyzed whether there was an enrichment for overlap with potential functional elements in our set of 756 DML related to COVID-19 compared to matched background DML using the experimentally-derived Functional element Overlap analysis of ReGions from EWAS tool (eFORGE) (Breeze et al., 2019). eFORGE uses 815 datasets from the ENCODE, Roadmap Epigenomics, and BLUEPRINT epigenomic mapping projects to detect enriched tissues, cell types, and genomic regions of DML from DNA methylation profiling studies. Our analysis utilizing the chromatin all 15-state marks reference revealed the greatest enrichment in actively transcribed genes of primary B cells (q value = 3.65e-10), mononuclear cells (q value = 4.39e-8), and neutrophils (q value = 1.03e-11) from peripheral blood suggesting potential cell composition and/or cell type-specific effects of COVID-19 infection in blood (**Supplementary Dataset 2**). In addition to these immune cell-type findings, we evaluated whether the observed epigenetic signature mimic methylation changes in SARS-CoV-2 target tissues. This analysis revealed significant enrichment in multiple organ systems supporting COVID-19 as a complex multisystem disorder (**Supplementary Dataset 2**). Notable enrichments were observed in actively transcribed genes of the digestive system (sigmoid colon, q value = 8.42e-8; duodenum smooth muscle, q value = 5.06e-8; rectal mucosa, q value = 3.19e-7; duodenum mucosa, q value = 3.19e-7), placenta (q value = 6.21e-06), spleen (q value = 1.25e-05, brain (anterior caudate, q value= 1.32e-05; hippocampus, q value= 2.13e-05), liver (q value = 3.92e-05), and lung (q value = 9.22e-05) (**Supplementary Dataset 2**). These data support the notion of COVID-19 impacting the epigenetic landscape as a multisystem disorder involving both immune cells and nonimmune cells in disease pathogenesis.

Among the top Δβ-value methylation changes comparing pre- and post-COVID-19 timepoints, we observed two differentially methylated loci (cg02037503 and cg23712970) in a gene promoter transcription start site regulatory region of the apoptotic chromatin condensation inducer 1 (*ACIN1*) gene **Supplementary Figure S2** that decreased in DNA methylation following COVID-19 by approximately 15% for both CpG sites (**Fig.1d,e**). This gene codes a nuclear protein that induces apoptotic chromatin condensation after activation by caspase-3 (Sahara et al., 1999), which we previously reported was increased in red blood cells of hospitalized COVID-19 participants (Plassmeyer et al., 2021). We also observed differential methylation at cg10846936 related to the caspase recruitment domain family member 14 (*CARD14*) gene that is involved in activating nuclear factor-kappa-B involved in immune inflammation. Participants’ methylation levels at this loci increased comparing pre- and post-COVID-19 timepoints from a mean of 44.47% to 53.44% (**Fig.1f**). Using a validation analysis of this loci in a public COVID-19 DNA methylation dataset GSE168739 (Castro de Moura et al., 2021), we observed that the mean methylation state of this loci in participants post-COVID-19 timepoint (mean DNAm = 53.5%) was more similar (*P* = 0.15, Tukey’s test) than participants pre-COVID-19 levels (mean DNAm = 44.6%) (*P* = 0.0001, Tukey’s test) compared to levels of this loci in a public blood DNA methylation dataset available from 407 confirmed COVID-19 participants (mean DNAm = 56.5%) (Castro de Moura et al., 2021) **Supplementary Figure S3**.

Additional top differentially methylated loci related to the corticotropin releasing hormone receptor 1 (*CRHR1,* cg09422970; **Fig.1g**), HIVEP Zinc Finger 3 (*HIVEP3,* cg11857452; **Fig.1h**), Meis Homeobox 2 (*MEIS2,* cg06471042; **Fig.1i**), Nuclear Receptor Coactivator 2 (*NCOA2,* cg20282780; **Fig.1j**), Progesterone Immunomodulatory Binding Factor 1 (*PIBF1,* cg00531853 and 15128396; **Fig.1k,l**), and Sterile Alpha Motif Domain Containing 4A (*SAMD4A,* cg18499294; **Fig.1m**) genes.

### 2.3 DNA methylation-based estimates of cell-type fractions in blood are significantly changed following COVID-19 infection

DNA methylation data can be utilized to infer fractions of immune cell types present in a heterogenous blood sample based on a reference list of CpGs identified from differentially methylated cell-types (Houseman et al., 2012). Hence, we used a paired t-test analysis and examined whether participants DNAm-based estimates of CD8+ T cells, CD4+ T cells, natural killer (NK) cells, B cells, monocytes, granulocytes, plasmablasts, exhausted T cells (CD8+CD28-CD45RA-), CD8 Naïve T cells and CD4 Naïve T cells significantly differed comparing pre- and post-COVID-19 timepoints. Since the human immune system undergoes dramatic aging-related changes, we stratified our analysis into two groups based on those under or over 50 years of age. We observed no significant differences in the inferred proportion of CD8+ T cells following COVID-19 for those under and over 50 years of age (**Fig.2a)**. Participants under 50 years of age showed significant increases in the percentage of CD4+ T cells in blood following COVID-19 (**Fig.2b)**. In contrast, in those over 50 years of age we observed a significant decrease in the percentage of CD4+ T cells in blood following COVID-19 (**Fig.2b)**, reflecting COVID-19 reports of lymphopenia. Additionally, we observed significant decreases in the percentage of B cells in those over 50 years of age (**Fig.2d)**. In contrast, we observed shifts in decreasing plasmablasts percentage (**Fig.2g)** and increasing CD4+ Naïve T cells were observed in those under 50 years of age following COVID-19 (**Fig.2j)**. Innate immune NK and monocytes cell proportions did not significantly change following COVID-19 (**Fig.2c,e)**. We also did not observe significant differences in granulocytes, exhausted CD8+ T cells, and CD8+ naïve T cells (**Fig.2f,h,i)**. Together, these findings suggest age-related COVID-19 shifts in specific immune cell types occur in healthy individuals.

**Figure 2.**
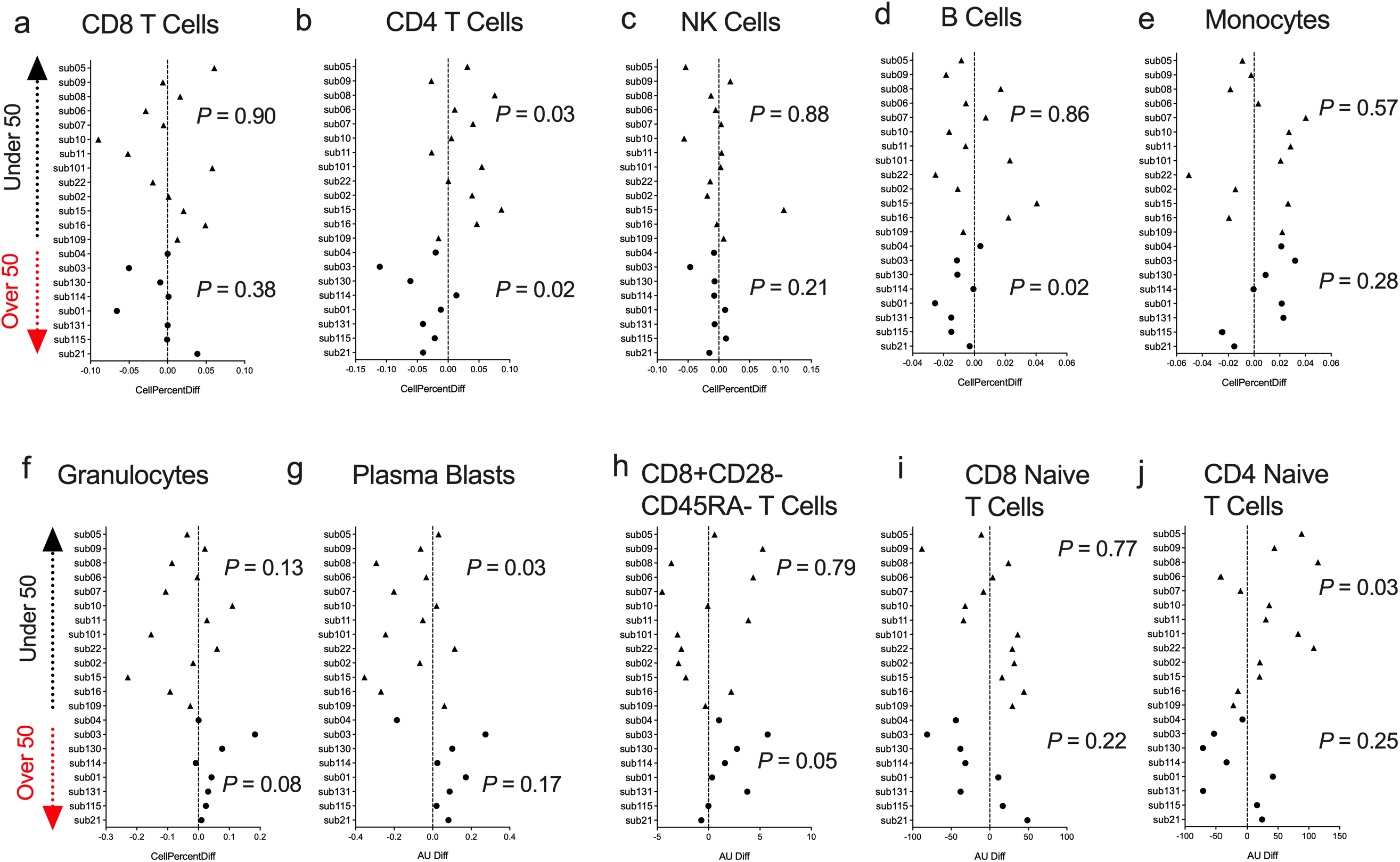
DNA methylation inferred blood immune cell type composition following mild/moderate COVID-19. **a-j.** Plots displaying the change in specific immune cell type populations inferred from DNA methylation in individuals pre- vs post-COVID-19 stratified by age. Triangles display participants under 50 years of age and circles display participants over 50 years of age.

We utilized the inferred cell type proportional changes following COVID-19 to examine relationships with age and the DNA methylation change pre- vs post-COVID-19 for ten of the top differentially methylated loci we had identified. This correlative analysis showed that chronological age was significantly associated with the percent change in DNA methylation for 9/10 DML, suggesting the signal of change for those DNA methylation sites related to age and potential plasticity loss (**Fig.3**). Moreover, chronological age was significantly associated with the inferred cell type proportional changes following COVID-19 for CD4+ T cells, plasmablasts, and CD4+ naïve T cells adding further support to our observed age-related COVID-19 shifts in specific immune cell types observations (**Fig.3**). Notably, we observed that shifts in inferred immune cell type proportions following COVID-19 significantly related to the extent of DNA methylation level changes for all of the DML we examined suggesting the COVID-19 DNA methylation signature to be substantially influenced by cell-type shifts (**Fig.3**).

**Figure 3.**
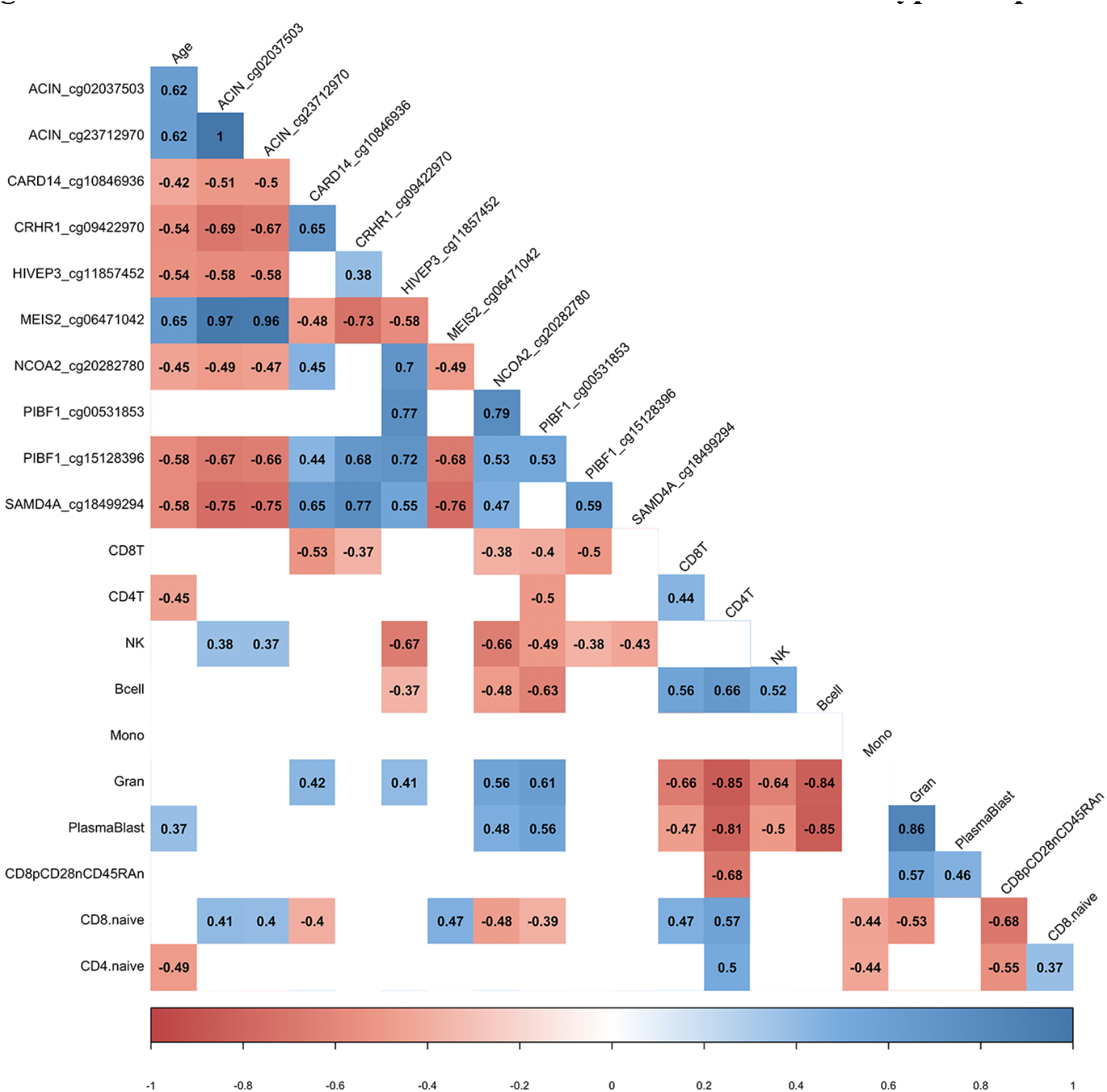
DML associated with COVID-19 relate to immune cell type composition. Correlogram plot of biological age, the change in DNA methylation levels for COVID-19 related DML, and the change in inferred immune cell type following COVID-19. Significant correlations displayed as solid box and Spearman’s rank correlation coefficient displayed.

### 2.4 DNA methylation changes in blood following COVID-19 overlap with COVID-19 related transcriptional gene sets

Various studies have identified transcriptional changes from SARS-CoV-2 infection(Blanco-Melo et al., 2020; Wilk et al., 2020). Since aberrant DNA methylation is commonly linked to transcriptional alterations, we sought to test whether the 516 genes containing a differentially methylated loci we identified through DNA methylation profiling of pre- and post-COVID-19 infection overlapped with COVID-19-related genes transcriptionally altered during SARS-CoV-2 infection utilizing the Enrichr COVID-19 gene set online tool (Kuleshov et al., 2016). This analysis revealed significant overlap with genes differentially expressed in SARS-CoV-2 animal models (Rhesus macaques blood, *P* = 0.017; mouse heart, *P* = 0.018; mouse spleen, *P* = 0.027; and hamster blood, *P* = 0.027), COVID-19 human biospecimens (late stage infection blood, *P* = 0.027; human cornea, *P* = 0.032), and *in vitro* infection models (Calu3, *P* = 0.030) (**Table 2**). These findings suggest DNA methylation patterns associated with COVID-19 likely play a role in transcriptional activation or repression of conserved host transcriptional responses to SARS-CoV-2 infection involving specific gene networks.

**Table 2:**
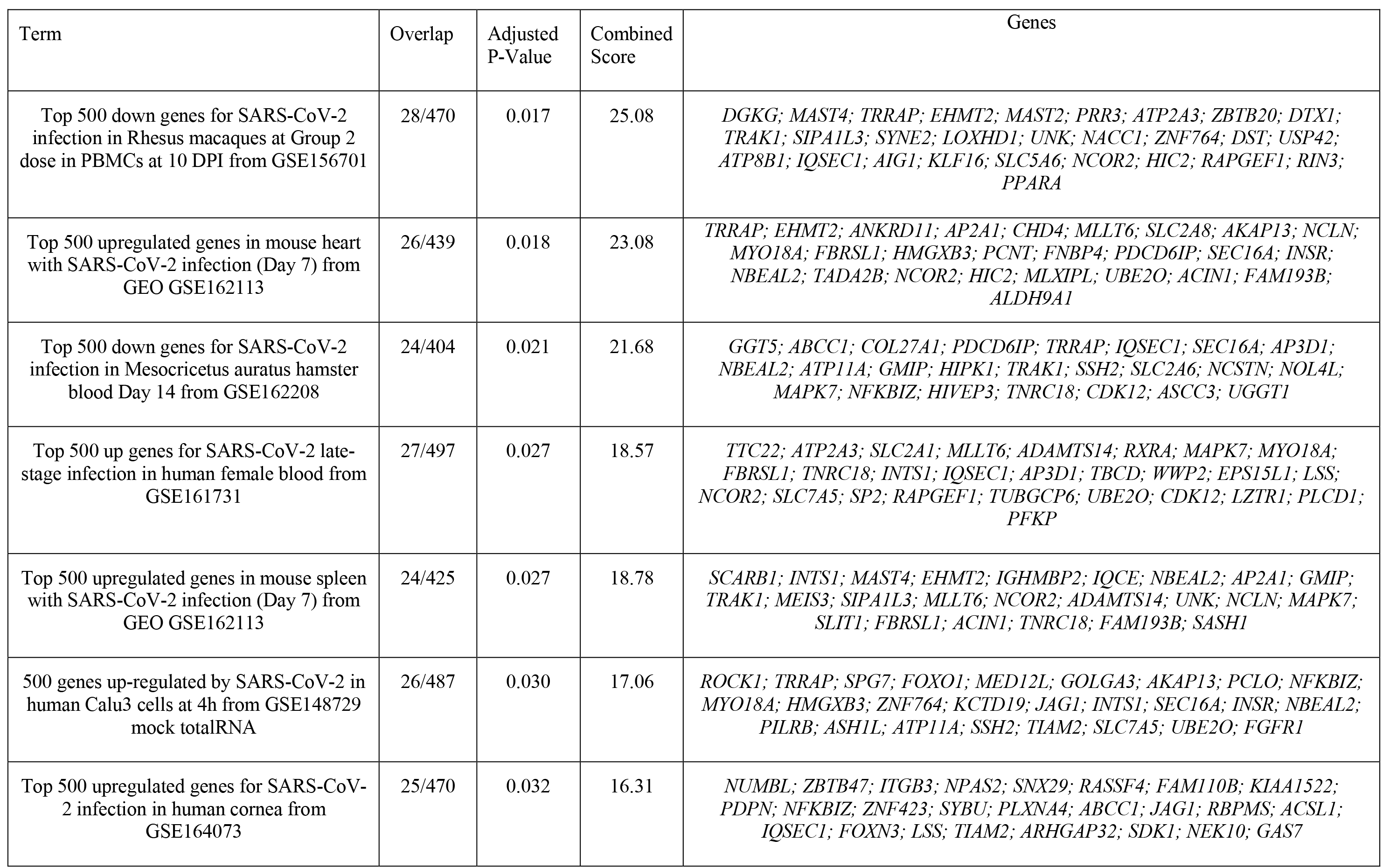
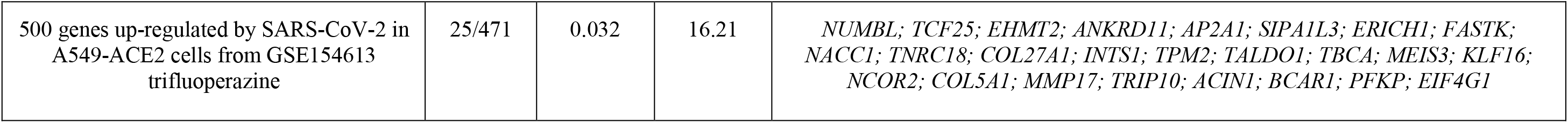
Differentially Methylated Loci Overlapping with Enrichr COVID-19 Related Gene Sets 2021.

### 2.5 Divergence in epigenetic clock estimates based on age related to COVID-19

Previous studies including our work reported epigenetic age perturbations associated with severe hospitalized COVID-19 in older individuals (Corley et al., 2021; Mongelli et al., 2021b). We sought to investigate in this pre- and post-COVID-19 cohort of non-hospitalized COVID-19 and relatively healthy individuals whether COVID-19 exposure impacted epigenetic clock estimates. We calculated epigenetic estimates for Horvath’s multi-tissue predictor DNAmAge based on 353 CpG sites (Horvath, 2013), the Horvath skin-and-blood clock based on 391 CpG sites (Horvath et al., 2018), Levine DNAmPhenoAge based on 513 CpG sites (Levine et al., 2018), Hannum’s clock based on 71 CpG sites (Hannum et al., 2013), the Lu telomere length predictor, and DNA methylation based mortality risk assessment (GrimAge (Lu et al., 2019)) using the Horvath online calculator. However, we found large bidirectional fluctuations in epigenetic age up to 8.99 years for DNAmAge, 4.49 years for Horvath skin-and-blood, 7.94 years for DNAmPheno Age, 6.25 years for Hannum’s clock, and 4.03 years for GrimAge that did not seem to be related to COVID-19 infection **Supplementary Figure S4**. Instead, this appeared to be attributable to widespread technical noise in DNAm measurement (Bose et al., 2014; Logue et al., 2017; Sugden et al., 2020), as previous studies have found that repeated measurements of the same sample to deviate up to 9 years (Higgins-Chen et al., 2021b). To mitigate the impacts of these unreliable epigenetic clock estimates, we applied a novel principal-component version of epigenetic clocks that permits a more reliable estimate for longitudinal studies (Higgins-Chen et al., 2021b). Next, we applied the principal-component epigenetic clocks algorithm based on 78,464 CpGs to the dataset to obtain PC-based epigenetic clock estimates and PC-based residuals after regressing PC-age predicted by the algorithm over chronological age for participants prior to and following COVID-19. Since age is a well-known risk factor for COVID-19 severity and our sample set contained participants that ranged in chronological age from 18 to 73 years, we stratified our longitudinal analysis of epigenetic clocks into two groups: people under 50 and people over 50 years of age. Application of our novel principal component-based computational solution to optimize the aging signal from epigenetic clocks and minimize noise revealed no significant differences in epigenetic age based on the PCHorvath1, PCHorvath 2, and PCHannum epigenetic clocks for both groups following COVID-19 (**Fig.4a-c**). Additionally, PCDNAmTL was not significantly altered following COVID-19 (**Fig.4d**). We observed that the PCPhenoAge clock was significantly increased in those over 50 years of age following COVID-19 by an average of 2.1 years (**Fig.4e,h**). In contrast to the observations for those over 50 years of age, PCPhenoAge was significantly decreased in those under 50 years of age following COVID-19 by an average of 2.06 years (**Fig.4g**). Chronological age significantly related to the extent of pre- vs post-COVID-19 epigenetic age increase in PCPhenoAge (**Fig.4i**). Moreover, we observed that PCGrimAge, a predictor of lifespan in unit of years was significantly increased in people over 50 years of age following COVID-19 by an average of 0.84 years (**Fig.4f,k**). Chronological age significantly related to the extent of pre- vs post-COVID-19 epigenetic age increase in PCGrimAge (**Fig.4l**). PCGrimAge was not significantly impacted in those under 50 years of age (**Fig.4j**).

**Figure 4.**
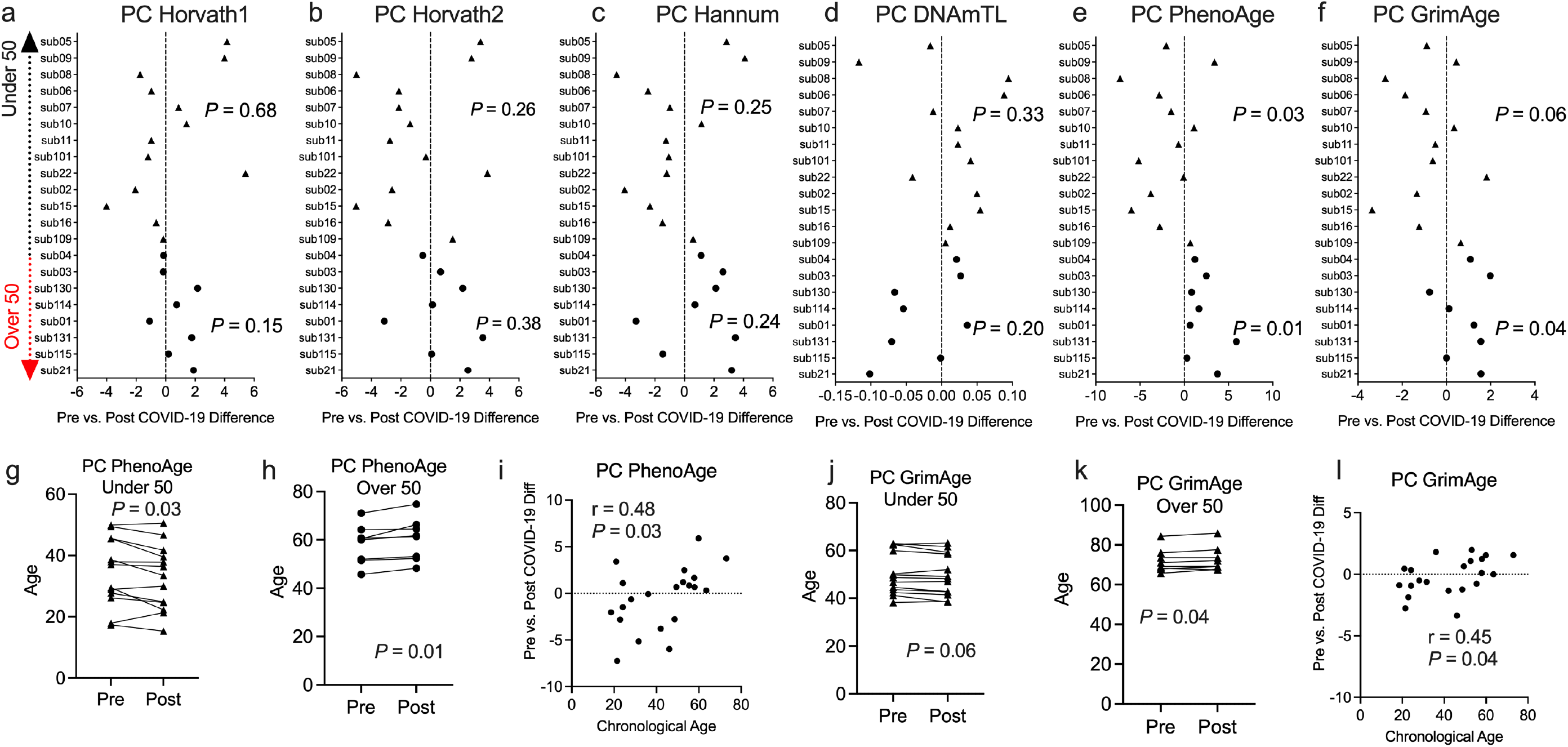
Divergence in principal component-based DNAmPhenoAge and GrimAge mortality risk increased based on age related to COVID-19. **a-f.** Plots displaying the change in principal component-based epigenetic clock age estimates in individuals pre- vs post-COVID-19 stratified by age. Triangles display participants under 50 years of age and circles display participants over 50 years of age. g-h. Plots displaying the change in principal component-based PhenoAge in individuals under and over 50 years of age pre- vs post-COVID-19. i. Correlation plot of chronological age and the change in PCPhenoAge pre- vs post-COVID-19. j-k. Plots displaying the change in principal component-based GrimAge in individuals under and over 50 years of age pre- vs post-COVID-19. l. Correlation plot of chronological age and the change in PCGrimAge pre- vs post-COVID-19.

We hypothesized that the COVID-19 associated change in PCPhenoAge and PCGrimAge was related to immune cell type compositional changes. Indeed, we observed that the increase in the PCPhenoAge clock estimate for participants following COVID-19 was significantly related to the magnitude of changes in the percent of CD4 T cells, B cells, Granulocytes, plasmablasts, exhausted T cells, CD8 naïve T cells, and CD4 naïve T cells (**Fig.5**). We did not observe significant relationships between the increase in PCPhenoAge and CD8T cells, NK cells, and monocytes (**Fig.5**). Moreover, we identified that the extent of increase in PCGrimAge estimates for participants following COVID-19 was significantly related to blood immune cell compositional changes in CD4 T cells, NK cells, B cells, granulocytes, and plasmablasts (**Fig.5**). The loss in percent CD4+ T cells following COVID-19 for all participants significantly related to older chronological age and increasing epigenetic age inferred from all epigenetic clocks (PCHorvath1, PCHorvath2, PCHannum, PCPhenoAge, and PCGrimAge) (**Fig.5**), supporting observations of lymphopenia related to COVID-19. Together, these findings suggest that the epigenetic aging signal related to COVID-19 exposure is driven by changes in blood immune cell type composition. We also examined whether COVID-19 impacted measures from a DNA methylation-based mitotic clock (“epiTOC” (Yang et al., 2016)) and quantification of the pace of biological aging (“DunedinPoAm” (Belsky et al., 2015, 2020)) and observed no significant differences pre vs post-COVID-19 in these measures (**Supplementary Figure S5**).

**Figure 5.**
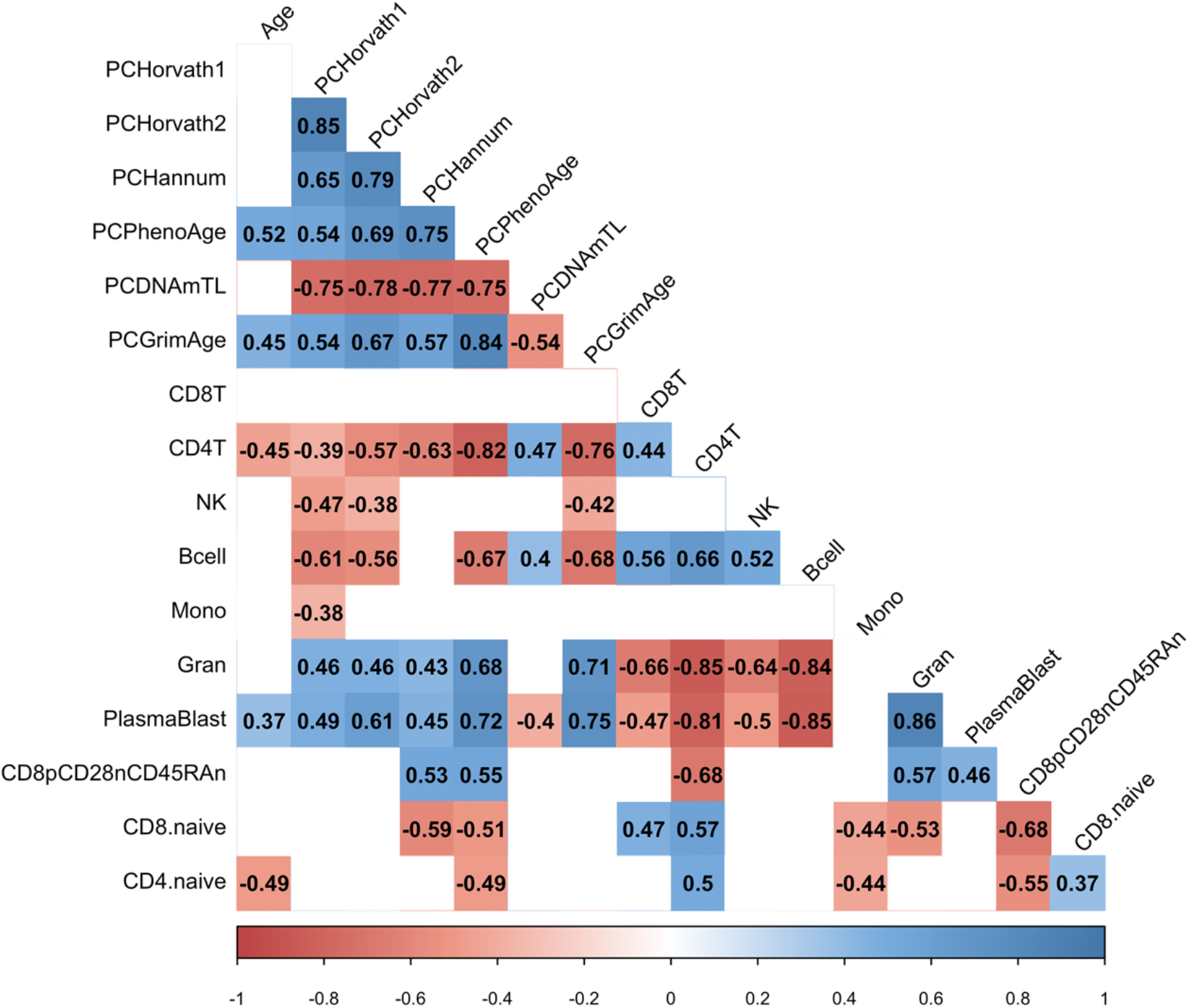
COVID-19 related epigenetic clock changes associate with immune cell type changes. Correlogram plot of biological age, the change in PC-based epigenetic clocks pre- vs. post-COVID-19, and the change in inferred immune cell type following COVID-19. Significant correlations displayed as solid box and Spearman’s rank correlation coefficient displayed.

### 2.6 mRNA COVID-19 vaccination in older individuals decreases epigenetic age and may reflect age-related B- cell and plasmablasts induction and expansion

To complement our pre- and post-COVID-19 exposure dataset, we sought to examine the impact of mRNA COVID-19 vaccination upon epigenetic clocks by obtaining DNA methylation profiles from blood of participants prior to and following mRNA COVID-19 vaccination. We examined 36 individuals (n=21 Females; n=15 Males) ranging in age from 22 to 69 years old that received either the Moderna (n=13) or Pfizer (n=23) mRNA vaccine. The median time since the second mRNA vaccine dose received by participants and DNA methylation data obtained for the post-vaccination timepoint was 57.9 days. We calculated principal-component epigenetic clock estimates and observed that PCHorvath1 and PCHorvath2 epigenetic age estimates were significantly decreased following complete mRNA vaccination comparing pre- and post-vaccination timepoints for all 36 participants by an average of 1.03 years and 1.58 years (**Fig.6a,b**). Exploratory analyses stratified by vaccine brand suggested that those over 50 years of age that received Moderna mRNA vaccination significantly reduced epigenetic age estimates based on PCHorvath1 by an average 2.75 years and PCHorvath2 by an average 3.91 years following complete vaccination (**Fig.7a,b**). In contrast, we observed no significant differences in epigenetic age estimates for people under 50 that received Moderna and for both those under and over 50 years of age that received Pfizer (**Fig.7a,b,g,h**). Whether these stratified results relate to Moderna vaccine containing a higher dose (100 micrograms) compared to Pfizer (30 micrograms) will need further examination. There was no significant difference in time from last dose and when DNA methylation data was obtained post-vaccination between vaccine brands (**Supplementary Figure S6**), suggesting time was not a confounding factor for vaccine differences in epigenetic age reduction in those over 50 years of age. In correlative analyses for all participants receiving an mRNA vaccine, we observed the extent of decreasing epigenetic age based on the PCHorvath1 and PCHorvath2 clocks and increasing PCDNAmTL significantly related to increasing chronological age (**Fig.7m,n,q**). We did not observe any significant differences from mRNA vaccination upon the PCHannum, PCPhenoAge, and PC DNAmTL(**Fig.7c,d,e,i,j,k**) and delta epigenetic age changes for PCHannum, PCPhenoAge, and PC GrimAge were not significantly related to chronological age (**Fig.7o,p,r**).

**Figure 6.**
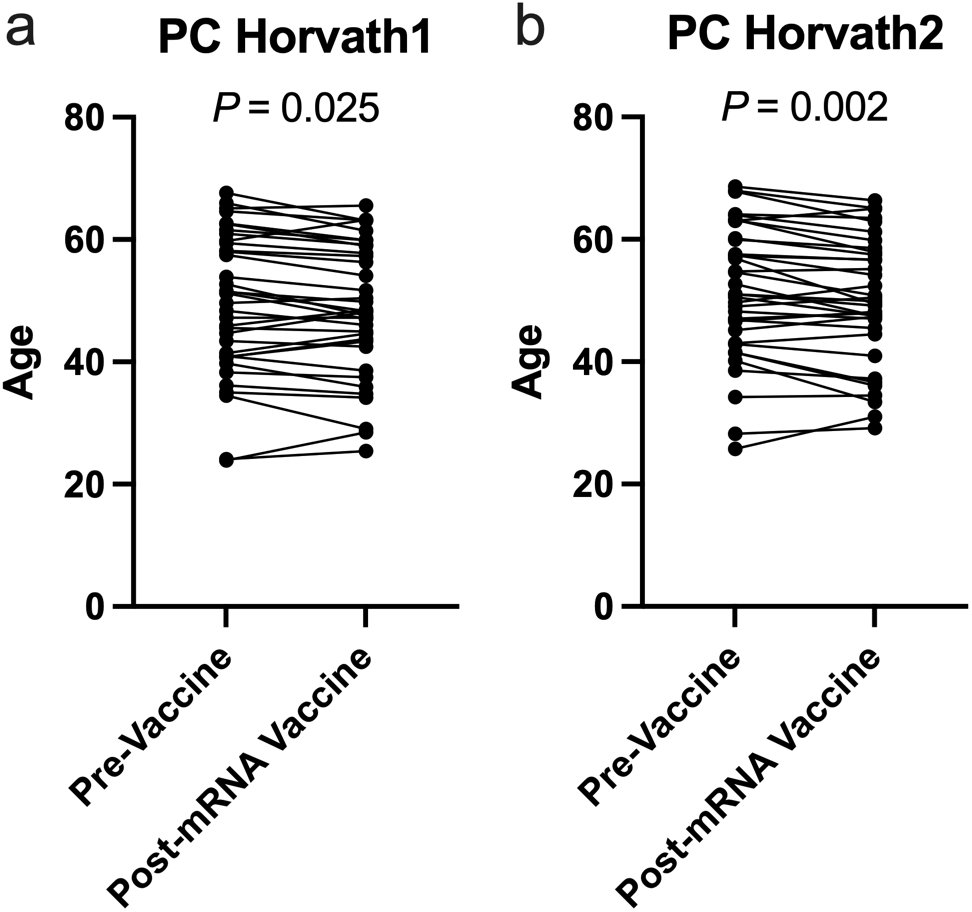
mRNA COVID-19 vaccination decreases PCHorvath1 and PCHorvath2 epigenetic age. **a.** Longitudinal plot of individuals PCHorvath1 and **b.** PCHorvath2 epigenetic age at pre- vaccine and post-mRNA vaccination time points. Paired t-test P value displayed.

**Figure 7.**
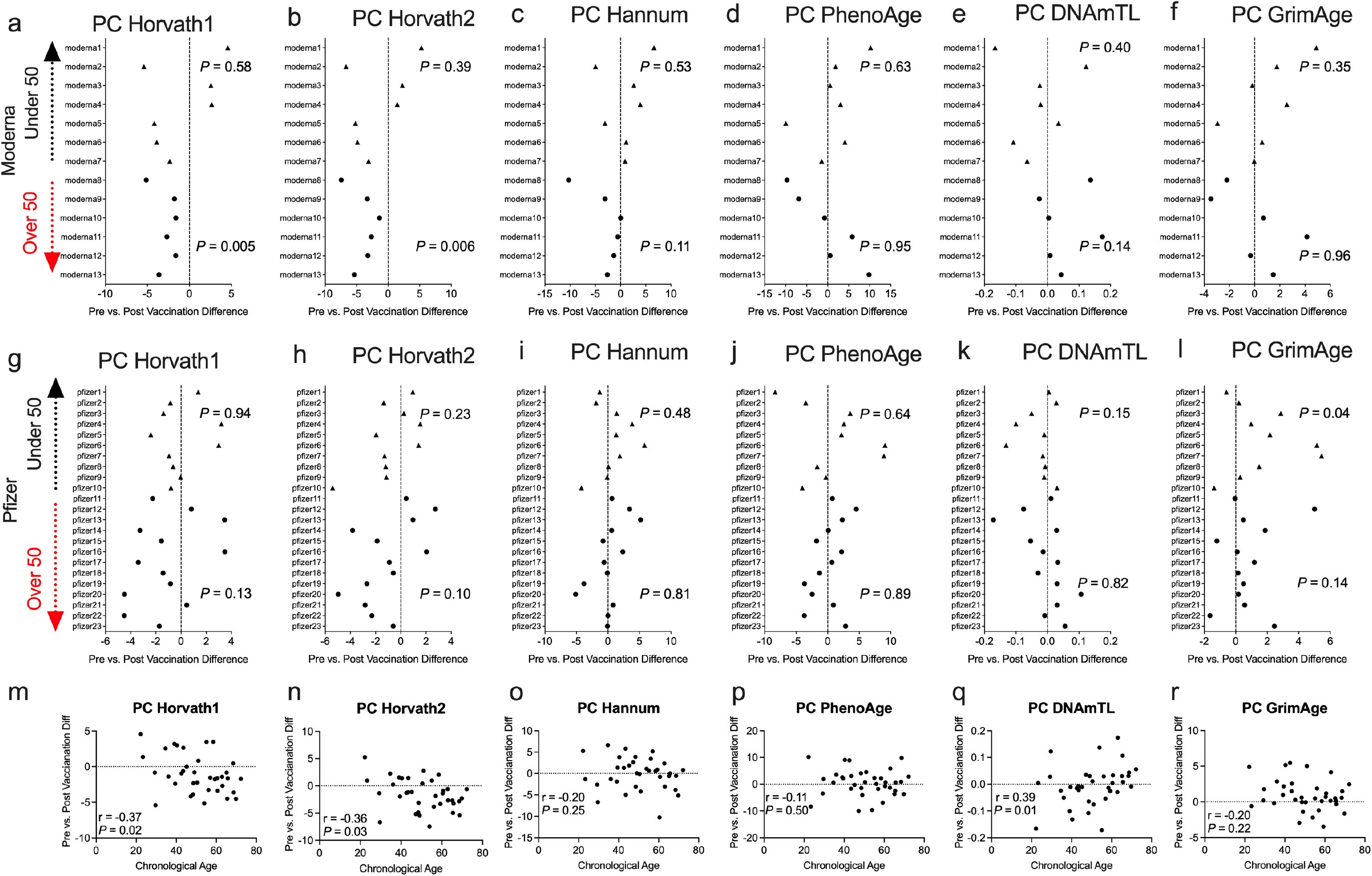
Moderna mRNA COVID-19 vaccination decreases principal component-based epigenetic age in individuals over 50. **a-f.** Plots displaying the change in principal component-based epigenetic clock age estimates in individuals pre- vs post-mRNA Moderna vaccination stratified by age. Triangles display participants under 50 years of age and circles display participants over 50 years of age. **g-i.** Plots displaying the change in principal component-based epigenetic clock age estimates in individuals pre- vs post-mRNA Pfizer vaccination stratified by age. **m-r.** Correlations between chronological age and pre- vs. post- mRNA vaccination change in PC-based epigenetic clock estimates.

Next, we examined whether the decrease in PCHorvath1 and PCHorvath2 following mRNA vaccination related to immune cell type compositional changes and/or DNA methylation inferred telomere length since previous data suggested telomere length related to influenza vaccine responses (Najarro et al., 2015). The delta change in PCHorvath1 following mRNA vaccination significantly associated with delta change in PCDNAmTL and plasmablasts cell type percentage following complete two dose mRNA vaccination (**Fig.8**). The delta change in PCHorvath2 following mRNA vaccination significantly associated with delta change in PCDNAmTL, B cell, granulocytes, and plasmablasts following complete two dose mRNA vaccination (**Fig.8**). Notably, we did not observe any significant relationships between the time elapsed from when participants received their second mRNA dose and post mRNA vaccine DNA methylation data was obtained and delta changes in all epigenetic clock estimates and cell type compositional changes following mRNA vaccination (**Fig.8**).

**Figure 8.**
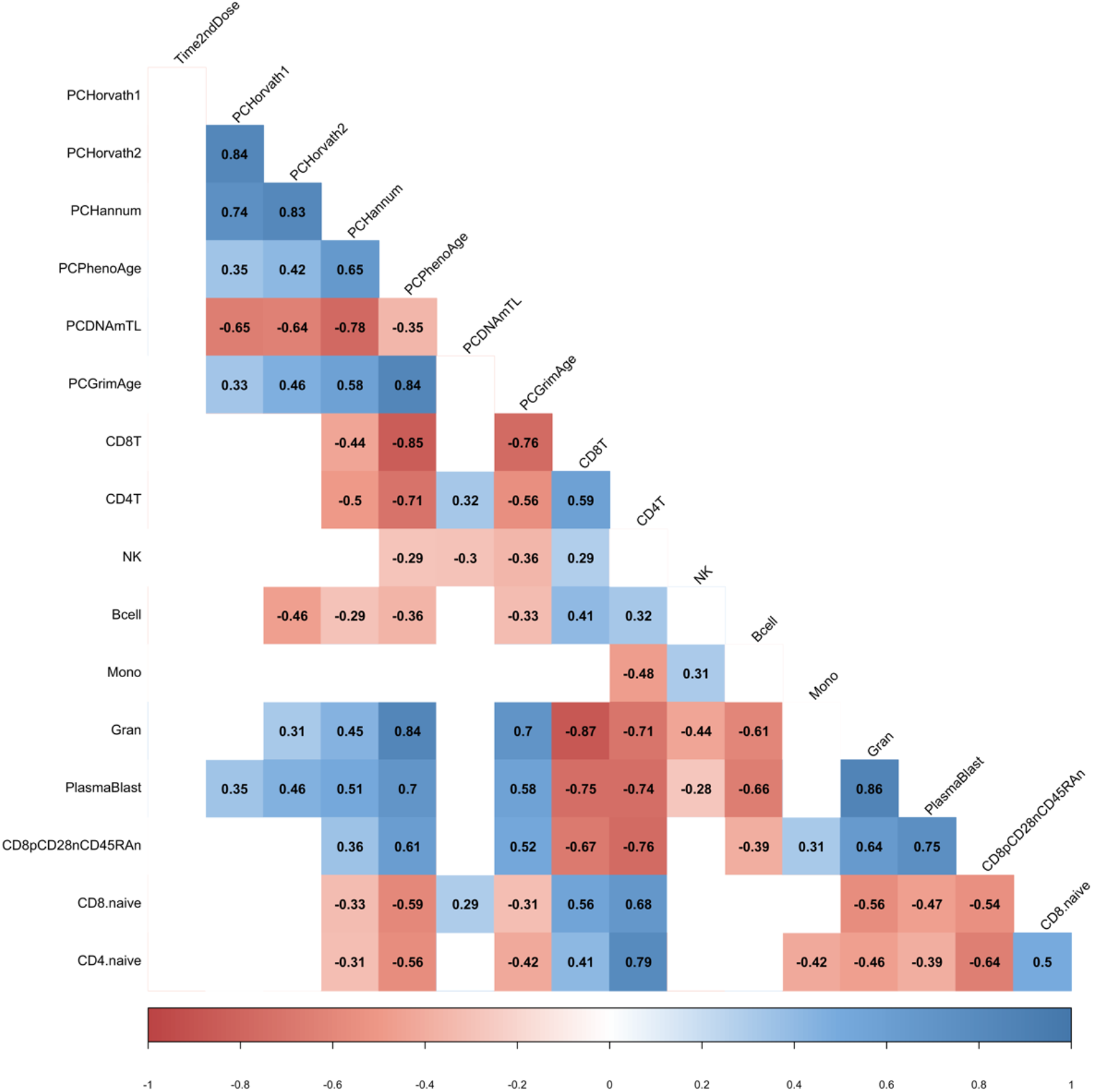
mRNA vaccine-related epigenetic clock changes associate with immune cell type changes. Correlogram plot of time since 2^nd^ dose, the change in PC-based epigenetic clocks pre- vs. post-COVID-19, and the change in inferred immune cell type following COVID-19. Significant correlations displayed as solid box and Spearman’s rank correlation coefficient displayed.

### 2.7 Short-term SARS-CoV-2 infection and exposure *in vitro* does not substantially impact epigenetic clocks

We tested whether artificial short-term *in vitro* exposure to SARS-CoV-2 virus (0.1 MOI) impacted PC-based epigenetic clocks in human peripheral mononuclear cells. We exposed viable PBMC’s from two uninfected donors (Donor 1, under 50 years of age; Donor 2: over 50 years of age) to a passage 4 stock of SARS-CoV-2 (0.1 MOI) *in vitro* for 60 hrs and compared DNA methylation levels to mock uninfected. We did not observe evidence of SARS-CoV-2 replication in PBMCs after 60 hrs as assessed by nucleocapsid flow cytometry assessments. PC-based epigenetic estimates of difference in epigenetic age comparing mock and SARS-CoV-2 exposed PBMC revealed changes less than 1 year for PCHorvath1, PCHorvath2, and PCHannum clocks (**Supplementary Figure S7**). Notably, we observed divergent donor-dependent impacts from addition of monophosphoryl lipid A (MPLA) stimulation, a TLR4 agonist, for 1 hr to 60hr SARS-CoV-2 exposed PBMC’s for all PC-based clocks (**Supplementary Figure S7**). As a comparator dataset, we generated DNA methylation data from mock and SARS-CoV-2 infected Calu-3 cells for 96 hours and observed epigenetic age did not increase for PC-based epigenetic clock comparing mock and infected cells.

## Discussion

In this pilot study, we examined whether SARS-CoV-2 infection and mRNA COVID-19 vaccination impacted DNA methylation states and epigenetic clocks in healthy individuals in the short term. Our findings revealed that significant differences in DNA methylation in blood associate with SARS-CoV-2 infection at 756 CpG sites, suggesting an immune cell-based epigenetic signature of COVID-19 may derive from aberrant DNA methylation states related to immune dysfunction induced by COVID-19. These findings support epigenetic findings from other groups that have reported distinct DNA methylation states in blood as a potential biomarker of COVID-19 (Balnis et al., 2021; Castro de Moura et al., 2021; Zhou et al., 2021). Moreover, our epigenetic clock findings reveal an age-related impact of epigenetic age increase associated with natural SARS-CoV-2 infection on the PCPhenoAge epigenetic clock and mortality risk estimate PCGrimAge in mild/moderate cases. Whether the extent and durability of this perturbation to these two epigenetic clock estimates is related to long COVID-19 or long-term aging outcomes remains an intriguing area for further investigation.

In contrast to natural SARS-CoV-2 infection, we observed that two epigenetic clocks (DNAmAge/PCHorvath1 and DNAmAgeSkinBlood/PCHorvath2) were decreased following mRNA COVID-19 vaccination in individuals over 50. The extent of decreased epigenetic age following mRNA-COVID-19 vaccination significantly related to changes in B cells and plasmablasts, highlighting the potential utility of epigenetic clocks in capturing vaccine responses and tracking the need for booster shots due to waning COVID-19 immunity in older individuals. These results are more robust because multiple clocks that putatively measure the same aging phenotype (i.e. PCGrimAge and PCPhenoAge predict mortality risk, while PCHorvath1 and PCHorvath2 track with chronological age) show similar relationships to COVID-19. Together, this pilot longitudinal epigenetic dataset of natural COVID-19 exposure and mRNA COVID-19 vaccination has important implications for research into the impact of COVID-19 on aging and the potential for mRNA vaccination to impact epigenetic aging in the immune system. Future research will need to examine whether COVID-19 and mRNA vaccine-related changes to epigenetic age are biologically meaningful. It is important to note that although epigenetic age robustly predicts age-related morbidity and mortality in cross-sectional studies (Horvath and Raj, 2018; Oblak et al., 2021), it is unknown if modifying epigenetic age in the short term leads to changes in long-term outcomes.

Prior research has shown that the host epigenetic landscape is altered during coronavirus infection (Schäfer and Baric, 2017). Evidence indicates that SARS-CoV-2 infection has a substantial impact upon the host immune cell epigenetic and transcriptional landscape in severe COVID-19 (Corley et al., 2021; Rendeiro et al., 2021). Our findings support a recent cross-sectional human DNA methylation study of COVID-19 that reported DNA methylation patterns of COVID-19 convalescents compared to uninfected controls (Huoman et al., 2021). In addition, our study provides the first examination of longitudinal DNA methylation changes in blood of healthy participants prior to and following test-confirmed mild/moderate COVID-19. We observed blood-based DNA methylation changes associated with COVID-19 exposure in healthy participants ranging in age with 261 differentially methylated CpGs identified. Among the COVID-19 differentially methylated loci we detected in blood, we observed hypermethylation related to the caspase recruitment domain family member 14 (*CARD14*) gene. This gene encodes a protein that has been shown to interact with BCL10 that functions as a positive regulator of cell apoptosis and NF-kappaB activation (Bertin et al., 2001). Moreover, CARD14 may play a role in protecting cells against apoptosis. We observed that the percent change in DNA methylation inferred immune cell type proportion for CD8 T cells for participants following COVID-19 exposure significantly associated with the DNA methylation percent change related to *CARD14*. This suggest that a subset of DNA methylation changes related to COVID-19 exposure were due to cell type compositional changes. Notably, we also observed the differentially methylated CpGs associated with COVID-19 were enriched in transcriptional gene sets identified from published SARS-CoV-2 human, animal model, and *in vitro* infection studies (Kuleshov et al., 2016, 2020). These findings suggest that DNA methylation changes associated with COVID-19 likely participate in the regulation and modulation of host gene expression from infection. Together, this first set of findings support the notion that distinct host DNA methylation states in circulating immune cells serves as a COVID-19 specific epigenetic signature. The durability of this COVID-19 epigenetic signature remains a key question for future study.

Recent work utilizing a cross sectional study design reported that epigenetic clocks are not altered in COVID-19 (Franzen et al., 2021). Our work contrasts with this report and suggests that specific epigenetic clocks may be altered by COVID-19 based on age. We utilized a more powerful longitudinal study design of individuals prior to and following test-confirmed COVID-19 and applied a novel principal component-based assessment of epigenetic clocks that mitigates issues with reliability in standard epigenetic clock algorithms. We observed a divergence in the epigenetic clock estimate PCPhenoAge and epigenetic clock mortality algorithm PCGrimAge based on age in individuals following COVID-19. Slight epigenetic age acceleration in the short term appeared in those individuals over 50 years of age that were infected with SARS-CoV-2. In contrast, epigenetic age appeared to reduce in those individuals under 50 years of age following COVID-19.

PCPhenoAge and PCGrimAge are among the strongest epigenetic predictors of mortality risk (Levine et al., 2018; Lu et al., 2019; Higgins-Chen et al., 2021a). These findings support the critical role of age as a COVID-19 risk factor and suggest that specific epigenetic clocks can capture an age-dependent perturbation to epigenetic clocks following COVID-19. Moreover, prior multi-omic analysis has shown that the Levine clock accelerates with cellular senescence and mitochondrial dysfunction (Liu et al., 2020). Prior studies of epigenetic clocks in COVID-19 utilized different sets of clocks which may explain their conflicting results (Corley et al., 2021; Franzen et al., 2021; Mongelli et al., 2021a). Interrogating a wide array of clocks simultaneously is essential for determining which clocks are most related to COVID-19 or vaccination. Furthermore, findings are more robust if multiple clocks predicting the same phenotype show the same relationship to COVID-19.

A plausible interpretation of the PCPhenoAge/PCGrimAge results after infection is an age-related signal of both immunosenescence and inflammaging. Once the immune system is activated in younger individuals by SARS-CoV-2 infection, they look younger by the epigenetic clock due to a robust activation of the immune response that reflects younger individuals (not that they are actually becoming younger). In older individuals, activation of non-specific inflammatory pathways after SARS-CoV-2 infection appears to increase epigenetic age because of the activation of pathways that overlap/are similar to inflammaging. An alternative interpretation for divergence in epigenetic age based on age during SARS-CoV-2 infection might involve the biological process of hormesis: moderate stressors can improve health by causing a compensatory response (Epel, 2020). COVID-19 might serve as a hormetic stress in non-hospitalized younger individuals, while it serves as a toxic stressor in older adults or any severe case.

Aging drives immunosenescence with implications for a decline in adaptive immunity resulting in reduced vaccine responses and vaccine durability in older adults. The age-related decline in immune function including reduced thymic output of naïve T cells and dampened B cell generation has notably led to decreased vaccine efficacy in older individuals (Soiza et al., 2021). Indeed, building evidence for COVID-19 indicates a declined humoral and cellular immune response in older individuals (Collier et al., 2021; Levin et al., 2021). Yet, failure to achieve a protected or durable response after vaccination is poorly understood despite occurring commonly among many elderly individuals. Our epigenetic clock data following mRNA COVID-19 vaccination revealed an age-related decrease in epigenetic age following vaccination. Our findings also revealed that the change in epigenetic age following vaccination was specifically related to immune-cell type compositional changes in the percentage of B cells, plasmablasts, and granulocytes. These findings support work showing that SARS-CoV-2 mRNA vaccines induce persistent germinal center B cell response that enables robust humoral immunity (Turner et al., 2021). Our findings do not provide any insights into the particular impact of the mRNA lipid nanoparticle compared to the expressed spike protein upon different epigenetic clocks. These compelling findings suggest that epigenetic profiles and specifically epigenetic clock estimates may provide insights into individual and age-related humoral immune responses to COVID-19 vaccination. Previous work has shown the impact of influenza vaccination on persistent epigenomic remodeling of immune cells (Wimmers et al., 2021) and explored the idea of whether epigenetic age could relate to vaccine responses in the context of influenza (Gensous et al., 2018).

Recent work examining T cell exhaustion after recovery from chronic infection in humans has found that epigenetic scars of CD8+ T cell exhaustion persists in humans (Yates et al., 2021), suggesting indelible imprints on the host immune cell epigenome from viral infection. The hypothesis of a persistent epigenetic dysregulation of host immune cells contributing to long-COVID-19 remains unclear. Whether durable changes to epigenetic clocks are reflected by epigenetic scars of particular immune cell types and relate to long-COVID-19 is a compelling hypothesis to pursue.

Our findings highlight the benefits of our computational solution using principal components for calculating PC-based epigenetic clocks for longitudinal studies (Higgins-Chen et al., 2021b). Using standard epigenetic clock estimates, we observed variation up to 9 years in participants pre- and post-timepoint samples epigenetic age estimates for all clocks that lead to non-significant results. While the application of PC-based epigenetic clocks pulled out a biological signal suggesting that mild/moderate COVID-19 from SARS-CoV-2 infection and mRNA vaccination impacted epigenetic clocks, the biological mechanisms that influence detrimental or beneficial changes in epigenetic clocks remains unclear. Ongoing research is deconstructing dissimilar epigenetic clocks and may provide further insights into the precise biological mechanisms captured from age-related alterations in the methylation landscape during infection and mRNA vaccination.

The developed mRNA vaccines for COVID-19 have been shown to elicit a potent humoral immune response and be highly efficacious at preventing COVID-19 and severe disease outcomes (Polack et al., 2020; Baden et al., 2021; Tartof et al., 2021). Our DNA methylation dataset was obtained at a median post second dose of around 2 months. Based on data showing durability of vaccine responses out to 6 months post-vaccination (Doria-Rose et al., 2021), our DNA methylation data was captured during an effective post-vaccine time frame window. Future studies will need to harness serial sample collection of participants during the course of mRNA vaccination and assess critical time points for impacts of mRNA vaccination upon epigenetic clocks. Moreover, our observations of age-related impacts from mRNA vaccination upon epigenetic age warrants further investigation to determine whether this measure may be relevant to age-related waning vaccine effectiveness. Lastly, epigenetic age assessments of participants that received heterologous prime-boost vaccination against COVID-19 (Borobia et al., 2021; Nordström et al., 2021; Pozzetto et al., 2021; Schmidt et al., 2021; Shaw et al., 2021) and heterologous booster vaccinations (Atmar et al., 2021) should be studied.

Considering the challenges with longitudinal blood collection and acquisition of epigenetic DNA methylation data from participants at timepoints prior to and following test-confirmed COVID-19, all of the published COVID-19 DNA methylation studies have been cross-sectional study designs (Bernardes et al., 2020; Balnis et al., 2021; Castro de Moura et al., 2021; Corley et al., 2021). Longitudinal epigenetic studies are considered the gold standard study design to mitigate interindividual variation in DNA methylation patterns and track environmental and pathogen-induced changes to the epigenome (Chen et al., 2018). Hence, our assessment of longitudinal DNA methylation of 21 participants provides a discovery dataset for examining the short-term impacts of COVID-19 upon the host immune cell epigenome and impact on epigenetic clock estimates. Our longitudinal COVID-19 DNA methylation dataset consisted of healthy participants that ranged across the lifespan from 18 to 73 years of age. Moreover, the collection of DNA methylation data following test-confirmed COVID-19 exposure occurred within a short-term time frame of a 6-month window and occurred early during the COVID-19 pandemic reflecting infection with a less evolved, less contagious, and potentially less severe SARS-CoV-2 virus than recent variants such as the Delta variant. Hence, our findings are limited by these potential factors. These longitudinal findings need to be confirmed in a larger sample size, across diverse regions and genotypes, among individuals across the lifespan, in people infected with emerging SARS-CoV-2 variants, and across COVID-19 severities including those individuals that recover and suffer from long-lasting symptoms termed post-COVID.

Despite the strengths of this longitudinal epigenetic study, there are several limitations. First, our longitudinal study design only included two time points to examine changes related to COVID-19 and mRNA vaccination comparing baseline and a short-term follow-up assessment of DNA methylation. future studies will need to study a larger sample size and determine whether these age-related divergent changes to epigenetic clocks are durable following COVID-19 and potentially relate to those with long**-**COVID-19 syndrome. Additionally, there was variation in the time following confirmed COVID-19 or mRNA COVID-19 vaccination for when the post sample assay for DNA methylation was completed. Yet, given the complete lack of longitudinal DNA methylation studies of COVID-19 and mRNA COVID-19 vaccination we provide discovery findings that are compelling regarding specific DNA methylation changes and epigenetic clocks that warrant further investigation. Future studies that have serial blood collection of participants throughout the course of mRNA- vaccination and even following booster shots will be extremely valuable for epigenetic clock investigations. Recent technological advancements based on Tagmentation-based Indexing of Methylation Sequencing (TIME-Seq) have scaled and reduced the cost of epigenetic age predictions permitting methodology for a more comprehensive study follow-up to our findings (Griffin et al., 2021). We also acknowledge the limited clinical data for participants and that the SARS-CoV-2 infection DNA methylation dataset may be relevant to a early genetic lineage not reflecting emerging variants being monitored nor variants of concern such as Delta (B.1.617.2) .

## 3 Methods

### 3.1 Study Cohort

Deidentified DNA methylation data was generated by TruDiagnostic as part of a retrospective non-randomized study to assess the effects of SARS-CoV-2 infection and mRNA vaccination upon DNA methylation patterns. Participants post-COVID-19 sample DNA methylation was test confirmed PCR testing or serology testing and occurred between August 2020 and March 2021. This study was approved by the Institute of Regenerative and Cellular Medicine IRB and Weill Cornell Medicine IRB.

### 3.2 DNA methylation Assessment

Peripheral whole blood was collected by lancet and capillary method into lysis buffer and DNA extracted. 500 ng of DNA were bisulfite converted using the EZ DNA Methylation kit (Zymo Research) according to the manufacturer’s instructions. Bisulfite-converted DNA samples were randomly assigned to a chip well on the Infinium HumanMethylationEPIC BeadChip, amplified, hybridized onto the array, stained, washed, and imaged with the Illumina iScan SQ instrument to obtain raw image intensities. DNA methylation data for longitudinal sampling of participant’s pre- and post-COVID-19 and pre- and post-vaccination time points was assayed for each participant at separate times.

### 3.3 DNA methylation analyses

Raw Methylation EPIC array IDAT intensity data was loaded and preprocessed in the R statistical programming language (http://www.r-project.org) using The Chip Analysis Methylation Pipeline (ChAMP, version 2.8.3)(Tian et al., 2017). IDAT files were loaded using the champ.load function. All samples passed quality control metrics. Comprehensive filtering was applied to the dataset for probes with detection P-values <0.01, all non-CpG probes, previously published SNP-related probes, multi-hit probes, and probes on sex chromosomes. Methylation beta-values ranging from 0 -1 (corresponding to unmethylated to methylated signal intensity) for each sample were normalized using the BMIQ function implemented in the ChAMP pipeline. DNA methylation epigenetic age parameters were calculated using Horvath’s web-based DNAm age calculator tool(Horvath, 2013; Lu et al., 2019). Cell type deconvolution estimates for blood were calculated using the EpiDISH package(Zheng et al., 2019). To identify differentially methylated loci, we utilized an FDR adjustment (Benjamini-Hochberg) and adjusted P value cutoff at 0.05 to compare participants pre-COVID-19 methylation data to post-COVID-19 methylation data. Genes related to differentially methylated loci were utilized for a COVID-19 gene set analyses from the Enrichr web tool(Kuleshov et al., 2016).

#### Epigenetic Clock Analysis and Estimating blood immune cell type composition

Epigenetic clock estimates, DNA methylation-based cell type deconvolution proportions, and epigenetic biomarkers were calculated using the online calculator (https://dnamage.genetics.ucla.new). Principal component-based epigenetic clock estimates were calculated utilizing an R script and 78,464 CpGs for each sample in a beta matrix. Mean imputation was utilized for missing values. Pace of aging was calculated utilizing the DunedinPACE algorithm (DunedinPoAm_45). To calculate pace of aging, noob-normalized and masked beta values were first processed from raw IDAT files using the Sesame R package(Zhou et al., 2018). To limit the number of batch effects caused by processing multiple bead chips, individual bead chips were processed at a time to generate the normalized beta values and then used to quantify pace of aging. The pace of aging metric was then calculated using the DunedinPACE algorithm described in Belsky et al. 2021(Belsky et al., 2021). Briefly, the algorithm uses 19 different physiological biomarkers to generate overall pace of aging from the Dunedin Study cohort (N = 1037). A standardized average rate of aging is then regressed using an elastic net regression model against methylation values generated from blood collected from the cohort at the age of 45, which identified 173 CpG sites that are associated with the pace of aging metric. The DunedinPACE algorithm was used to calculate the pace of aging measure obtained from authors. Analyses were performed in R 4.1.1 and RStudio Version 1.4.1717. Figures were made using GraphPad and corrplot R package.

##### *In vitro* SARS-CoV-2 infection and exposure

SARS-CoV-2 virus (isolate USA-WA1/2020 (BEI resources; NR-52281) was propagated and titrated in Vero E6 cell lines. De-identified donor PBMC specimens were obtained from Astarte Biological for *in vitro* exposure to 0.1 MOI SARS-CoV-2 for 60 hrs. Calu-3 cells were infected for 96 hrs.

## 7 Author Contributions

MJC, LNC, RS conceived of and designed the study. MJC, APS, ATH wrote the manuscript. FC, IR, CA, HW, TM, VD performed recruitment, data collection, analyses, and DNA methylation experiments. MEL and ATH provided PC-Clocks algorithm. MS performed *in vitro* SARS-CoV-2 experiments.

## Data Availability

All data produced in the present study are available upon reasonable request to the authors

## Acknowledgments

We gratefully acknowledge the study participants and their physicians who made this work possible.

## 12 Data Availability Statement

The datasets generated for this study can be found in the NIH GEO database. GEO Submission (Pending)

## References

Atmar, R. L., Lyke, K. E., Deming, M. E., Jackson, L. A., Branche, A. R., El Sahly, H. M., et al. (2021). Heterologous SARS-CoV-2 Booster Vaccinations - Preliminary Report. medRxiv. doi:10.1101/2021.10.10.21264827.

Baden, L. R., El Sahly, H. M., Essink, B., Kotloff, K., Frey, S., Novak, R., et al. (2021). Efficacy and Safety of the mRNA-1273 SARS-CoV-2 Vaccine. N. Engl. J. Med. 384, 403–416.

Balnis, J., Madrid, A., Hogan, K. J., Drake, L. A., Chieng, H. C., Tiwari, A., et al. (2021). Blood DNA methylation and COVID-19 outcomes. Clin. Epigenetics 13, 118.

Belsky, D. W., Caspi, A., Arseneault, L., Baccarelli, A., Corcoran, D. L., Gao, X., et al. (2020). Quantification of the pace of biological aging in humans through a blood test, the DunedinPoAm DNA methylation algorithm. Elife 9. doi:10.7554/eLife.54870.

Belsky, D. W., Caspi, A., Corcoran, D. L., Sugden, K., Poulton, R., Arseneault, L., et al. (2021). Quantification of the pace of biological aging in humans through a blood test: the DunedinPACE DNA methylation algorithm. bioRxiv. doi:10.1101/2021.08.30.21262858.

Belsky, D. W., Caspi, A., Houts, R., Cohen, H. J., Corcoran, D. L., Danese, A., et al. (2015). Quantification of biological aging in young adults. Proc. Natl. Acad. Sci. U. S. A. 112, E4104–10.

Bernardes, J. P., Mishra, N., Tran, F., Bahmer, T., Best, L., Blase, J. I., et al. (2020). Longitudinal Multi-omics Analyses Identify Responses of Megakaryocytes, Erythroid Cells, and Plasmablasts as Hallmarks of Severe COVID-19. Immunity 53, 1296–1314.e9.

Bertin, J., Wang, L., Guo, Y., Jacobson, M. D., Poyet, J.-L., Srinivasula, S. M., et al. (2001). CARD11 and CARD14 Are Novel Caspase Recruitment Domain (CARD)/Membrane-associated Guanylate Kinase (MAGUK) Family Members that Interact with BCL10 and Activate NF-κB*. J. Biol. Chem. 276, 11877–11882.

Blanco-Melo, D., Nilsson-Payant, B. E., Liu, W.-C., Uhl, S., Hoagland, D., Møller, R., et al. (2020). Imbalanced Host Response to SARS-CoV-2 Drives Development of COVID-19. Cell 181, 1036–1045.e9.

Borobia, A. M., Carcas, A. J., Pérez-Olmeda, M., Castaño, L., Bertran, M. J., García-Pérez, J., et al. (2021). Immunogenicity and reactogenicity of BNT162b2 booster in ChAdOx1-S-primed participants (CombiVacS): a multicentre, open-label, randomised, controlled, phase 2 trial. Lancet 398, 121–130.

Bose, M., Wu, C., Pankow, J. S., Demerath, E. W., Bressler, J., Fornage, M., et al. (2014). Evaluation of microarray-based DNA methylation measurement using technical replicates: the Atherosclerosis Risk In Communities (ARIC) Study. BMC Bioinformatics 15, 312.

Boulias, K., Lieberman, J., and Greer, E. L. (2016). An Epigenetic Clock Measures Accelerated Aging in Treated HIV Infection. Mol. Cell 62, 153–155.

Breeze, C. E., Reynolds, A. P., van Dongen, J., Dunham, I., Lazar, J., Neph, S., et al. (2019). eFORGE v2.0: updated analysis of cell type-specific signal in epigenomic data. Bioinformatics 35, 4767–4769.

Castro de Moura, M., Davalos, V., Planas-Serra, L., Alvarez-Errico, D., Arribas, C., Ruiz, M., et al. (2021). Epigenome-wide association study of COVID-19 severity with respiratory failure. EBioMedicine 66, 103339.

Chen, R., Xia, L., Tu, K., Duan, M., Kukurba, K., Li-Pook-Than, J., et al. (2018). Longitudinal personal DNA methylome dynamics in a human with a chronic condition. Nat. Med. doi:10.1038/s41591-018-0237-x.

Collier, D. A., Ferreira, I. A. T. M., Kotagiri, P., Datir, R. P., Lim, E. Y., Touizer, E., et al. (2021). Age-related immune response heterogeneity to SARS-CoV-2 vaccine BNT162b2. Nature 596, 417–422.

Corley, M. J., Pang, A. P. S., Dody, K., Mudd, P. A., Patterson, B. K., Seethamraju, H., et al. (2021). Genome-wide DNA methylation profiling of peripheral blood reveals an epigenetic signature associated with severe COVID-19. J. Leukoc. Biol. doi:10.1002/JLB.5HI0720-466R.

Doria-Rose, N., Suthar, M. S., Makowski, M., O’Connell, S., McDermott, A. B., Flach, B., et al. (2021). Antibody Persistence through 6 Months after the Second Dose of mRNA-1273 Vaccine for Covid-19. N. Engl. J. Med. 384, 2259–2261.

Epel, E. S. (2020). The geroscience agenda: Toxic stress, hormetic stress, and the rate of aging. Ageing Res. Rev. 63, 101167.

Franzen, J., Nüchtern, S., Tharmapalan, V., Vieri, M., Nikolić, M., Han, Y., et al. (2021). Epigenetic Clocks Are Not Accelerated in COVID-19 Patients. Int. J. Mol. Sci. 22. doi:10.3390/ijms22179306.

Gensous, N., Franceschi, C., Blomberg, B. B., Pirazzini, C., Ravaioli, F., Gentilini, D., et al. (2018). Responders and non-responders to influenza vaccination: A DNA methylation approach on blood cells. Exp. Gerontol. 105, 94–100.

Gómez-Díaz, E., Jordà, M., Peinado, M. A., and Rivero, A. (2012). Epigenetics of host-pathogen interactions: the road ahead and the road behind. PLoS Pathog. 8, e1003007.

Griffin, P. T., Kane, A. E., Trapp, A., Li, J., McNamara, M. S., Meer, M. V., et al. (2021). Ultra-cheap and scalable epigenetic age predictions with TIME-Seq. bioRxiv, 2021.10.25.465725. doi:10.1101/2021.10.25.465725.

Hannum, G., Guinney, J., Zhao, L., Zhang, L., Hughes, G., Sadda, S., et al. (2013). Genome-wide methylation profiles reveal quantitative views of human aging rates. Mol. Cell 49, 359–367.

Higgins-Chen, A. T., Thrush, K. L., and Levine, M. E. (2021a). Aging biomarkers and the brain. Semin. Cell Dev. Biol. 116, 180–193.

Higgins-Chen, A. T., Thrush, K. L., Wang, Y., Kuo, P.-L., Wang, M., Minteer, C. J., et al. (2021b). A computational solution for bolstering reliability of epigenetic clocks: Implications for clinical trials and longitudinal tracking. bioRxiv, 2021.04.16.440205. doi:10.1101/2021.04.16.440205.

Hillary, R. F., Stevenson, A. J., Cox, S. R., McCartney, D. L., Harris, S. E., Seeboth, A., et al. (2021). An epigenetic predictor of death captures multi-modal measures of brain health. Mol. Psychiatry 26, 3806–3816.

Hillary, R. F., Stevenson, A. J., McCartney, D. L., Campbell, A., Walker, R. M., Howard, D. M., et al. (2020). Epigenetic measures of ageing predict the prevalence and incidence of leading causes of death and disease burden. Clin. Epigenetics 12, 115.

Horvath, S. (2013). DNA methylation age of human tissues and cell types. Genome Biol. 14, R115.

Horvath, S., and Levine, A. J. (2015). HIV-1 Infection Accelerates Age According to the Epigenetic Clock. J. Infect. Dis. 212, 1563–1573.

Horvath, S., Oshima, J., Martin, G. M., Lu, A. T., Quach, A., Cohen, H., et al. (2018). Epigenetic clock for skin and blood cells applied to Hutchinson Gilford Progeria Syndrome and ex vivo studies. Aging 10, 1758–1775.

Horvath, S., and Raj, K. (2018). DNA methylation-based biomarkers and the epigenetic clock theory of ageing. Nat. Rev. Genet. 19, 371–384.

Houseman, E. A., Accomando, W. P., Koestler, D. C., Christensen, B. C., Marsit, C. J., Nelson, H. H., et al. (2012). DNA methylation arrays as surrogate measures of cell mixture distribution. BMC Bioinformatics 13, 86.

Huoman, J., Sayyab, S., Apostolou, E., Karlsson, L., Porcile, L., Rizwan, M., et al. (2021). Mild SARS-CoV-2 infection modifies DNA methylation of peripheral blood mononuclear cells from COVID-19 convalescents. bioRxiv. doi:10.1101/2021.07.05.21260014.

Kuleshov, M. V., Clarke, D. J. B., Kropiwnicki, E., Jagodnik, K. M., Bartal, A., Evangelista, J. E., et al. (2020). The COVID-19 Gene and Drug Set Library. Res Sq. doi:10.21203/rs.3.rs-28582/v1.

Kuleshov, M. V., Jones, M. R., Rouillard, A. D., Fernandez, N. F., Duan, Q., Wang, Z., et al. (2016). Enrichr: a comprehensive gene set enrichment analysis web server 2016 update. Nucleic Acids Res. 44, W90–7.

Kuo, C.-L., Pilling, L. C., Atkins, J. C., Masoli, J., Delgado, J., Tignanelli, C., et al. (2020). COVID-19 severity is predicted by earlier evidence of accelerated aging. medRxiv. doi:10.1101/2020.07.10.20147777.

Levin, E. G., Lustig, Y., Cohen, C., Fluss, R., Indenbaum, V., Amit, S., et al. (2021). Waning Immune Humoral Response to BNT162b2 Covid-19 Vaccine over 6 Months. N. Engl. J. Med. doi:10.1056/NEJMoa2114583.

Levine, M. E., Lu, A. T., Quach, A., Chen, B. H., Assimes, T. L., Bandinelli, S., et al. (2018). An epigenetic biomarker of aging for lifespan and healthspan. Aging 10, 573–591.

Liu, Z., Leung, D., Thrush, K., Zhao, W., Ratliff, S., Tanaka, T., et al. (2020). Underlying features of epigenetic aging clocks in vivo and in vitro. Aging Cell 19, e13229.

Logue, M. W., Smith, A. K., Wolf, E. J., Maniates, H., Stone, A., Schichman, S. A., et al. (2017). The correlation of methylation levels measured using Illumina 450K and EPIC BeadChips in blood samples. Epigenomics 9, 1363–1371.

Lu, A. T., Quach, A., Wilson, J. G., Reiner, A. P., Aviv, A., Raj, K., et al. (2019). DNA methylation GrimAge strongly predicts lifespan and healthspan. Aging 11, 303–327.

Mongelli, A., Barbi, V., Gottardi Zamperla, M., Atlante, S., Forleo, L., Nesta, M., et al. (2021a). Evidence for Biological Age Acceleration and Telomere Shortening in COVID-19 Survivors. Int. J. Mol. Sci. 22. doi:10.3390/ijms22116151.

Mongelli, A., Barbi, V., Zamperla, M. G., Atlante, S., Forleo, L., Nesta, M., et al. (2021b). Evidence for biological age acceleration and telomere shortening in covid-19 survivors. bioRxiv. doi:10.1101/2021.04.23.21255973.

Morales-Nebreda, L., McLafferty, F. S., and Singer, B. D. (2019). DNA methylation as a transcriptional regulator of the immune system. Transl. Res. 204, 1–18.

Mueller, A. L., McNamara, M. S., and Sinclair, D. A. (2020). Why does COVID-19 disproportionately affect older people? Aging 12, 9959–9981.

Najarro, K., Nguyen, H., Chen, G., Xu, M., Alcorta, S., Yao, X., et al. (2015). Telomere Length as an Indicator of the Robustness of B- and T-Cell Response to Influenza in Older Adults. J. Infect. Dis. 212, 1261–1269.

Nordström, P., Ballin, M., and Nordström, A. (2021). Effectiveness of heterologous ChAdOx1 nCoV-19 and mRNA prime-boost vaccination against symptomatic Covid-19 infection in Sweden: A nationwide cohort study. Lancet Reg Health Eur, 100249.

Oblak, L., van der Zaag, J., Higgins-Chen, A. T., Levine, M. E., and Boks, M. P. (2021). A systematic review of biological, social and environmental factors associated with epigenetic clock acceleration. Ageing Res. Rev. 69, 101348.

Pidsley, R., Zotenko, E., Peters, T. J., Lawrence, M. G., Risbridger, G. P., Molloy, P., et al. (2016). Critical evaluation of the Illumina MethylationEPIC BeadChip microarray for whole-genome DNA methylation profiling. Genome Biol. 17, 208.

Plassmeyer, M., Alpan, O., Corley, M. J., Premeaux, T. A., Lillard, K., Coatney, P., et al. (2021). Caspases and therapeutic potential of caspase inhibitors in moderate-severe SARS CoV2 infection and long COVID. Allergy. doi:10.1111/all.14907.

Polack, F. P., Thomas, S. J., Kitchin, N., Absalon, J., Gurtman, A., Lockhart, S., et al. (2020). Safety and Efficacy of the BNT162b2 mRNA Covid-19 Vaccine. N. Engl. J. Med. 383, 2603–2615.

Pozzetto, B., Legros, V., Djebali, S., Barateau, V., Guibert, N., Villard, M., et al. (2021). Immunogenicity and efficacy of heterologous ChadOx1/BNT162b2 vaccination. Nature. doi:10.1038/s41586-021-04120-y.

Reiterer, M., Rajan, M., Gómez-Banoy, N., Lau, J. D., Gomez-Escobar, L. G., Ma, L., et al. (2021). Hyperglycemia in acute COVID-19 is characterized by insulin resistance and adipose tissue infectivity by SARS-CoV-2. Cell Metab. 33, 2174–2188.e5.

Rendeiro, A. F., Ravichandran, H., Bram, Y., Chandar, V., Kim, J., Meydan, C., et al. (2021). The spatial landscape of lung pathology during COVID-19 progression. Nature. doi:10.1038/s41586-021-03475-6.

Sahara, S., Aoto, M., Eguchi, Y., Imamoto, N., Yoneda, Y., and Tsujimoto, Y. (1999). Acinus is a caspase-3-activated protein required for apoptotic chromatin condensation. Nature 401, 168– 173.

Schäfer, A., and Baric, R. S. (2017). Epigenetic Landscape during Coronavirus Infection. Pathogens 6. doi:10.3390/pathogens6010008.

Schmidt, T., Klemis, V., Schub, D., Schneitler, S., Reichert, M. C., Wilkens, H., et al. (2021). Cellular immunity predominates over humoral immunity after homologous and heterologous mRNA and vector-based COVID-19 vaccine regimens in solid organ transplant recipients. Am. J. Transplant. doi:10.1111/ajt.16818.

Shaw, R. H., Stuart, A., Greenland, M., Liu, X., Nguyen Van-Tam, J. S., Snape, M. D., et al. (2021). Heterologous prime-boost COVID-19 vaccination: initial reactogenicity data. Lancet 397, 2043–2046.

Soiza, R. L., Scicluna, C., and Thomson, E. C. (2021). Efficacy and safety of COVID-19 vaccines in older people. Age Ageing 50, 279–283.

Sugden, K., Hannon, E. J., Arseneault, L., Belsky, D. W., Corcoran, D. L., Fisher, H. L., et al. (2020). Patterns of Reliability: Assessing the Reproducibility and Integrity of DNA Methylation Measurement. Patterns (N Y) 1. doi:10.1016/j.patter.2020.100014.

Tartof, S. Y., Slezak, J. M., Fischer, H., Hong, V., Ackerson, B. K., Ranasinghe, O. N., et al. (2021). Effectiveness of mRNA BNT162b2 COVID-19 vaccine up to 6 months in a large integrated health system in the USA: a retrospective cohort study. Lancet. doi:10.1016/S0140-6736(21)02183-8.

Tian, Y., Morris, T. J., Webster, A. P., Yang, Z., Beck, S., Feber, A., et al. (2017). ChAMP: updated methylation analysis pipeline for Illumina BeadChips. Bioinformatics 33, 3982–3984.

Turner, J. S., O’Halloran, J. A., Kalaidina, E., Kim, W., Schmitz, A. J., Zhou, J. Q., et al. (2021). SARS-CoV-2 mRNA vaccines induce persistent human germinal centre responses. Nature 596, 109–113.

Wilk, A. J., Rustagi, A., Zhao, N. Q., Roque, J., Martínez-Colón, G. J., McKechnie, J. L., et al. (2020). A single-cell atlas of the peripheral immune response in patients with severe COVID-19. Nat. Med. 26, 1070–1076.

Wimmers, F., Donato, M., Kuo, A., Ashuach, T., Gupta, S., Li, C., et al. (2021). The single-cell epigenomic and transcriptional landscape of immunity to influenza vaccination. Cell 184, 3915–3935.e21.

Yang, Z., Wong, A., Kuh, D., Paul, D. S., Rakyan, V. K., Leslie, R. D., et al. (2016). Correlation of an epigenetic mitotic clock with cancer risk. Genome Biol. 17, 205.

Yates, K. B., Tonnerre, P., Martin, G. E., Gerdemann, U., Al Abosy, R., Comstock, D. E., et al. (2021). Epigenetic scars of CD8+ T cell exhaustion persist after cure of chronic infection in humans. Nat. Immunol. 22, 1020–1029.

Zheng, S. C., Breeze, C. E., Beck, S., Dong, D., Zhu, T., Ma, L., et al. (2019). EpiDISH web server: Epigenetic Dissection of Intra-Sample-Heterogeneity with online GUI. Bioinformatics. doi:10.1093/bioinformatics/btz833.

Zhou, S., Zhang, J., Xu, J., Zhang, F., Li, P., He, Y., et al. (2021). An epigenome-wide DNA methylation study of patients with COVID-19. Ann. Hum. Genet. doi:10.1111/ahg.12440.

Zhou, W., Triche, T. J., Jr, Laird, P. W., and Shen, H. (2018). SeSAMe: reducing artifactual detection of DNA methylation by Infinium BeadChips in genomic deletions. Nucleic Acids Res. 46, e123.

