## Supplemental Materials for "Longitudinal study of DNA methylation and epigenetic clocks prior to and following test-confirmed COVID-19 and mRNA vaccination"

SUPPLEMENTARY MATERIALS

**Alina PS Pang[1#], Albert T. Higgins-Chen[2,3#], Florence Comite[4,5], Ioana Raica[4], Christopher Arboleda[4], Hannah Went[6], Tavis Mendez[6], Michael Schotsaert[7,8], Varun Dwaraka[6], Ryan Smith[6], Morgan E. Levine[9], Lishomwa Ndhlovu[1], Michael J. Corley[1]***

[1] Division of Infectious Diseases, Department of Medicine, Weill Cornell Medicine, New York, NY, USA

[2]Department of Psychiatry, Yale University School of Medicine, New Haven, CT, USA

[3]VA Connecticut Healthcare System, West Haven, CT, USA

[4]Comite Center for Precision Medicine & Health, New York, NY, USA

[5]Lenox Hill Hospital/Northwell, New York, NY, USA

[6]TruDiagnostic, Lexington, KY, USA

[7]Department of Microbiology, Icahn School of Medicine at Mount Sinai, New York, NY, USA

[8]Global Health and Emerging Pathogens Institute, Icahn School of Medicine at Mount Sinai, New York, NY, USA

[9]Department of Pathology, Yale University School of Medicine, New Haven, CT, USA

**#Authors Contributed Equally**

**Supplementary Figure S1.** Enrichr gene enrichment showing top biological processes and KEGG pathways for genes associated with differentially methylated loci.

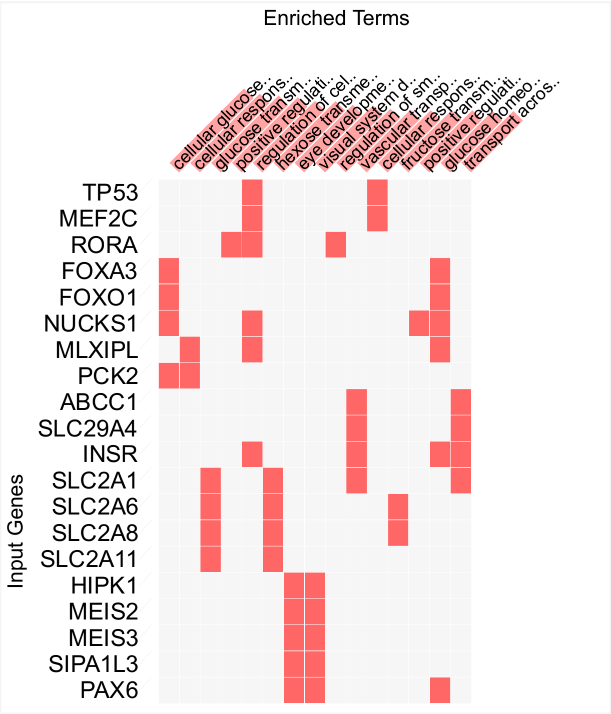


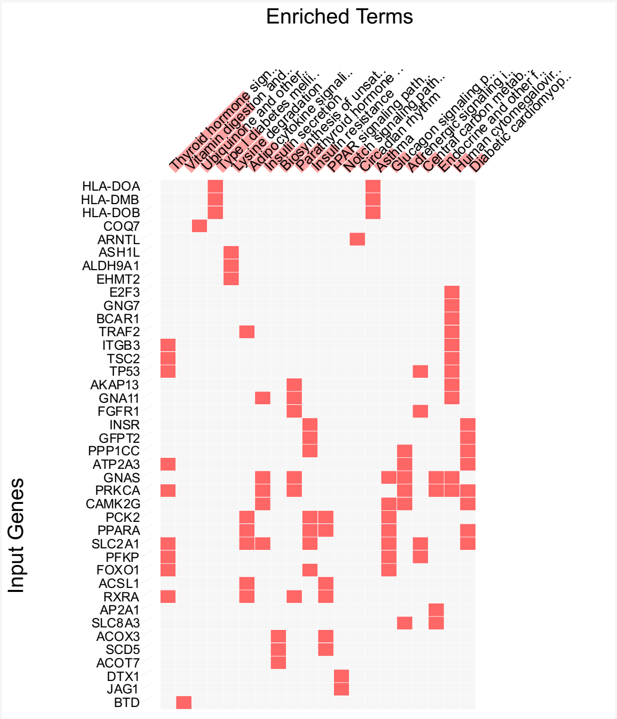

**Supplementary Figure S2.** Gene promoter transcription start site regulatory region of the apoptotic chromatin condensation inducer 1 (*ACIN1*)

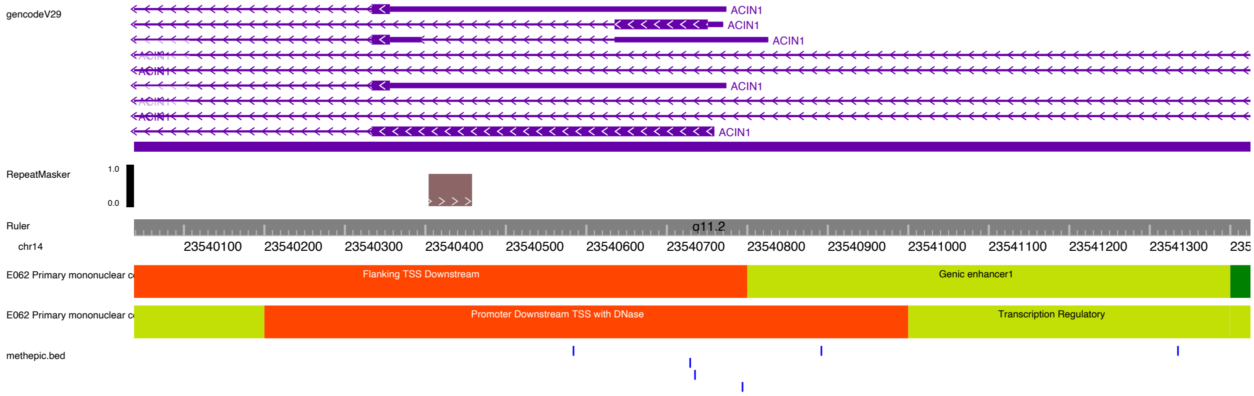

**Supplementary Figure S3.** Validation of cg10846936 in blood from 407 COVID-19 participants obtained from GEO GSE168739.

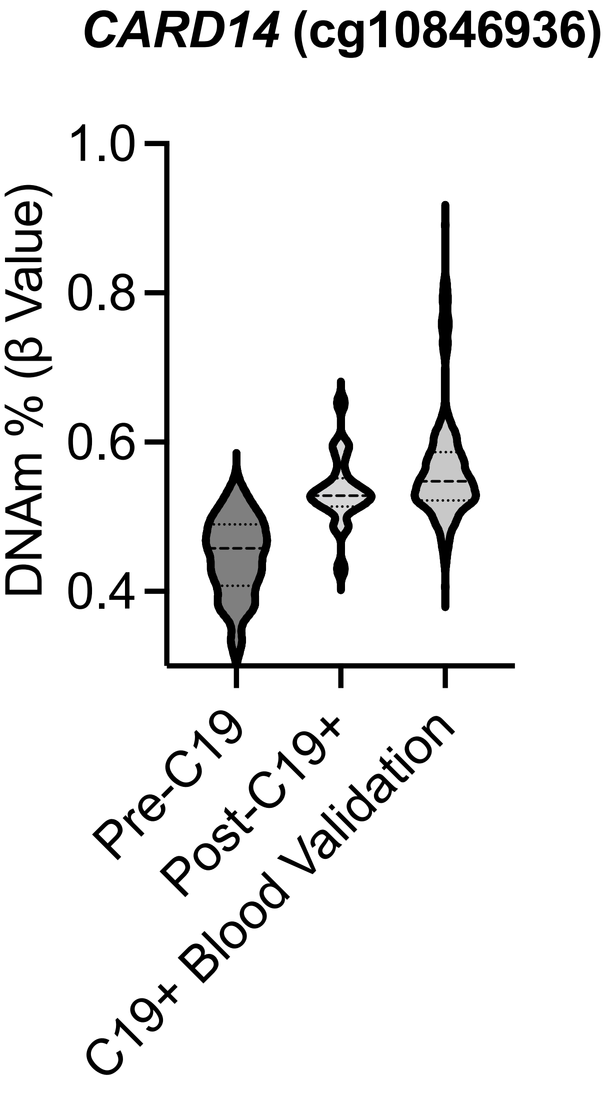

### Supplementary Figure S4

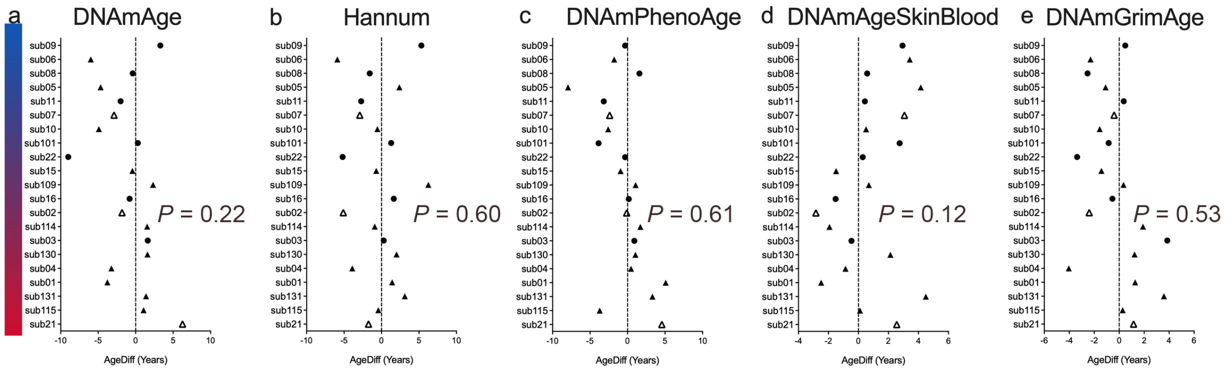

**Supplementary Figure S5**

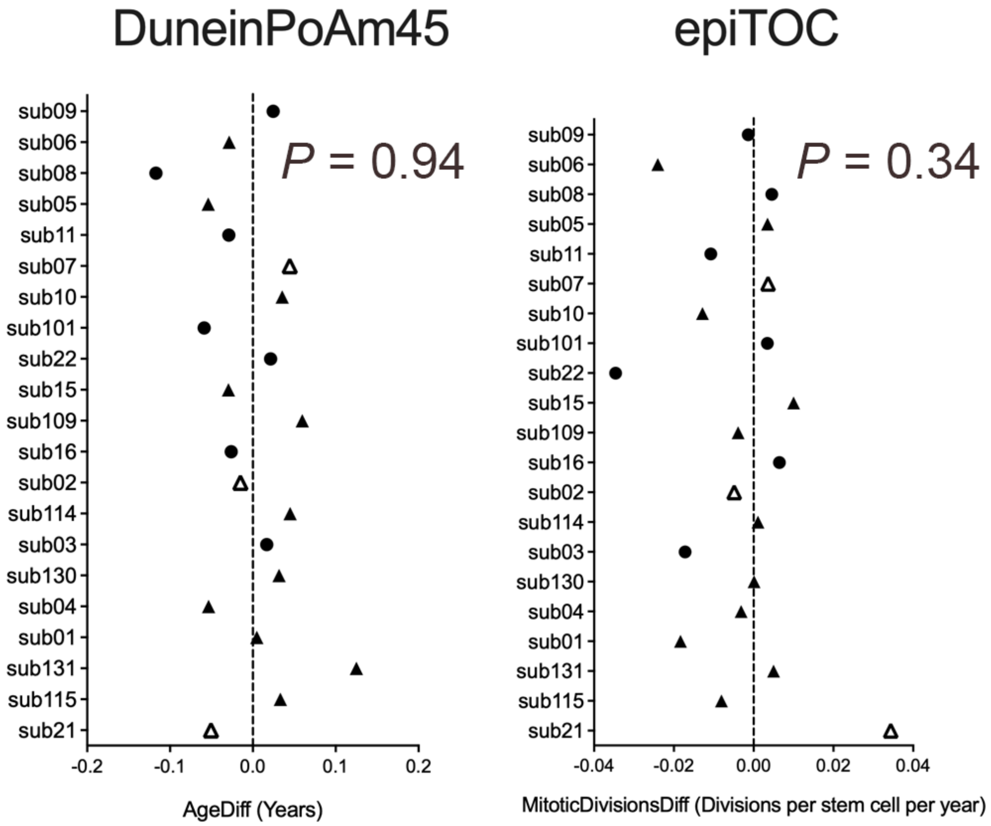

**Supplementary Figure S6.**

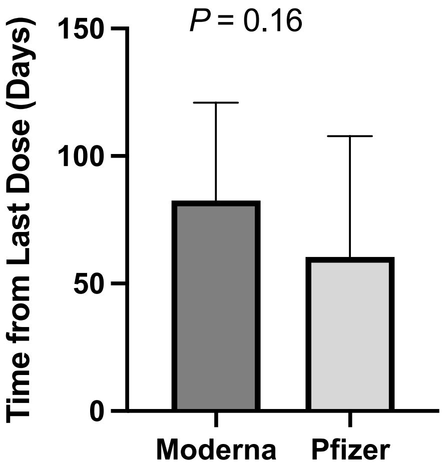

**Supplementary Figure S7.**

PC-based epigenetic age estimates in Calu-3 and Human Donor PBMC exposed to SARS-CoV-2 *in vitro*.

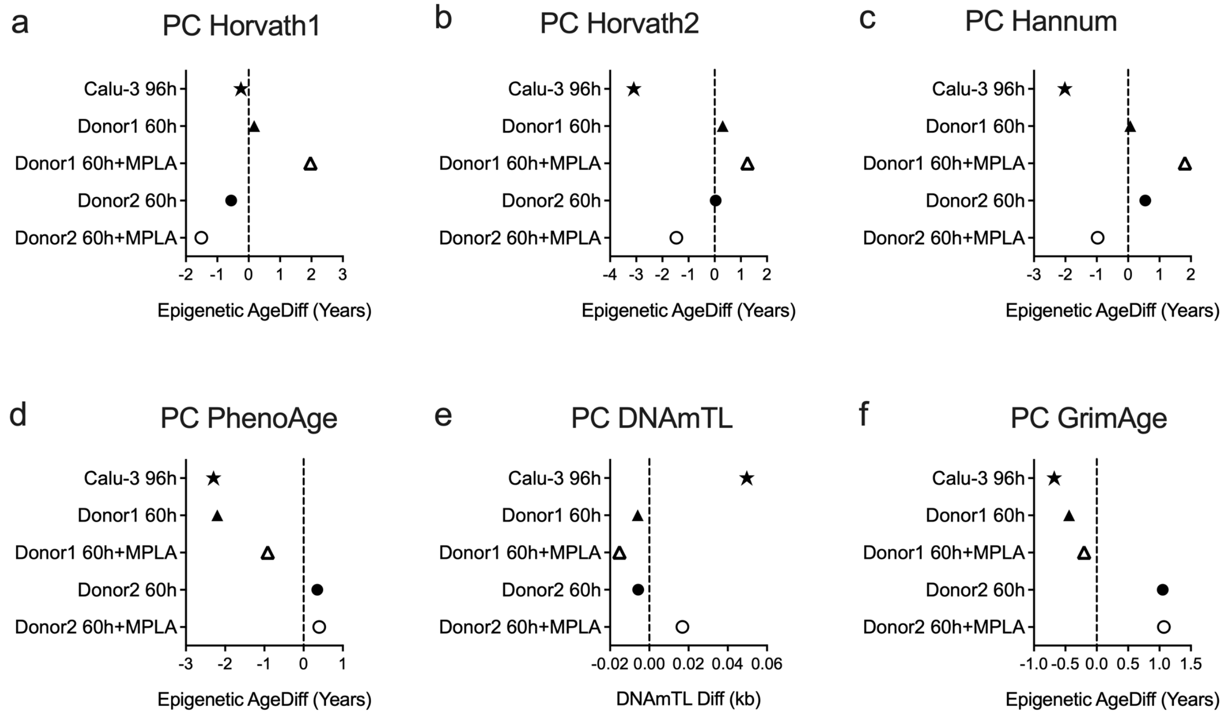

**Supplementary Figure S8.**
Validation of epigenetic age estimates in blood from participants at pre- and post-COVID-19 timepoints.

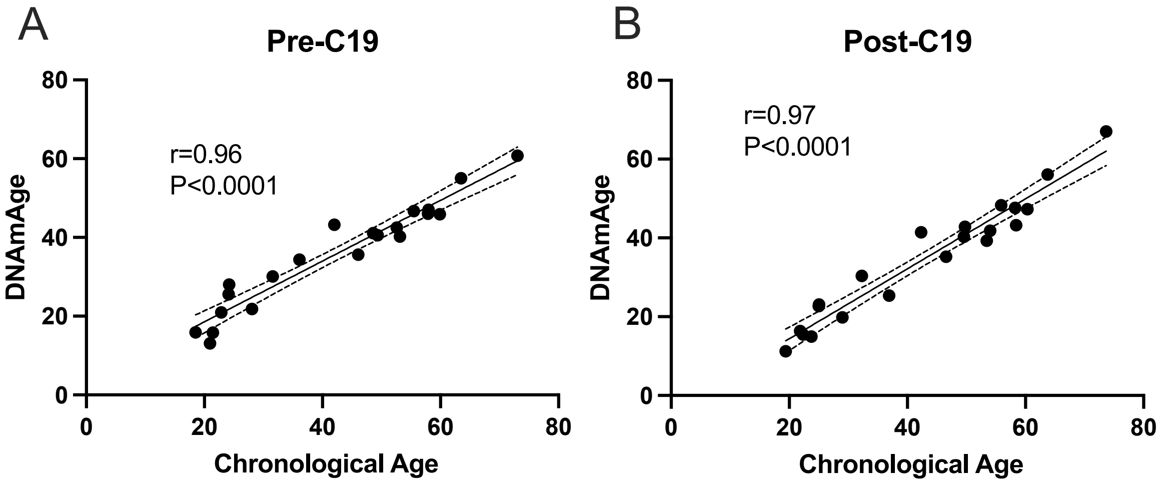
